# Prognostic Clinical and Biological Markers for Amyotrophic Lateral Sclerosis Disease Progression: Validation and Implications for Clinical Trial Design and Analysis

**DOI:** 10.1101/2024.08.12.24311876

**Authors:** Michael Benatar, Eric A Macklin, Andrea Malaspina, Mary-Louise Rogers, Eran Hornstein, Vittoria Lombardi, Danielle Renfrey, Stephanie Shepheard, Iddo Magen, Yahel Cohen, Volkan Granit, Jeffrey M Statland, Jeannine M Heckmann, Rosa Rademakers, Caroline A McHutchison, Leonard Petrucelli, Corey T McMillan, Joanne Wuu, the CReATe Consortium PGB1 Study Investigators

**Affiliations:** Department of Neurology, University of Miami Miller School of Medicine, Miami, FL, USA; Departments of Neurology and Medicine, Massachusetts General Hospital, Harvard Medical School, Boston, MA USA; UCL Queen Square Motor Neuron Disease Center, UCL Queen Square Institute of Neurology, University College London, Queen Square, London, UK; Flinders Health and Medical Research Institute, College of Medicine and Public Health, Flinders University, Adelaide, South Australia, Australia; Department of Molecular Genetics and Molecular Neuroscience, Weizmann Institute of Science, Israel; Department of Neurology, University of Kansas Medical Center, Kansas City, KS USA; Division of Neurology, Department of Medicine, University of Cape Town, South Africa; VIB Center for Molecular Neurology, Department of Biomedical Sciences, University of Antwerp, Antwerp, Belgium; Department of Neuroscience, Mayo Clinic, Jacksonville, Florida, USA; School of Philosophy, Psychology, and Language Sciences, The University of Edinburgh, Edinburgh, UK; Euan MacDonald Centre for Motor Neuron Disease Research, The University of Edinburgh, Edinburgh, UK; Department of Neurology, University of Pennsylvania Perelman School of Medicine, Philadelphia, PA, USA

**Author notes:** **<u>Correspondence to</u>:** Michael Benatar MD, PhD, or Joanne Wuu, ScM, Department of Neurology, University of Miami Miller School of Medicine, 1120 NW 14 Street, CRB Suite 1300, Miami, FL, 33136, USA.; or.

**Keywords:** Prognostic biomarkers, Context-of-use, ALS clinical trials, Neurofilament

## Abstract

**Background:** With increasing recognition of the value of incorporating prognostic markers into amyotrophic lateral sclerosis (ALS) trial design and analysis plans, there is a pressing need to understand *which* among the prevailing clinical and biochemical markers have real value, and *how* they can be optimally used.

**Methods:** A subset of patients with ALS recruited through the multi-center Phenotype-Genotype-Biomarker study (clinicaltrials.gov: NCT02327845) was identified as “trial-like” based on meeting common trial eligibility criteria. Clinical phenotyping was performed by evaluators trained in relevant assessments. Serum neurofilament light (NfL) and phosphorylated neurofilament heavy (pNfH), urinary p75^ECD^, plasma microRNA-181, and an array of biochemical and clinical measures were evaluated for their prognostic value. Associations with functional progression were estimated by random-slopes mixed models of ALS functional rating scale-revised (ALSFRS-R) score. Associations with survival were estimated by log-rank test and Cox proportional hazards regression. Potential sample size savings from adjusting for given biomarkers in a hypothetical trial were estimated.

**Findings:** Baseline serum NfL is a powerful prognostic biomarker, predicting survival and ALSFRS-R rate of decline. Serum NfL <40pg/ml and >100pg/ml correspond to future ALSFRS-R slopes of ∼0.5 and 1.5 points/month, respectively. Serum NfL also adds value to the best available clinical predictors, encapsulated by the European Network to Cure ALS (ENCALS) predictor score. In models of functional decline, the addition of NfL yields ∼25% sample size saving above those achieved by inclusion of either clinical predictors or ENCALS score alone. The prognostic value of serum pNfH, urinary p75^ECD^, and plasma miR-181ab is more limited.

**Interpretation:** Among the multitude of biomarkers considered, only blood NfL adds value to the ENCALS prediction model and should be incorporated into analysis plans for all ongoing and future ALS trials. Defined thresholds of NfL might also be used in trial design, for enrichment or stratified randomisation, to improve trial efficiency.

**Funding:** NIH (U01-NS107027, U54-NS092091). ALSA (16-TACL-242).

**Research in Context:** *Evidence Before This Study:* The phenotypic heterogeneity of ALS poses a challenge for clinical trials, making it more difficult to discern therapeutic effects of investigational agents amidst the noise of natural variability. Prognostic markers are important tools to help mitigate this issue. A host of clinical markers and putative biomarkers have been proposed to have prognostic value, but their relative utility, especially when considered jointly, and the practical implications of their use, have not been well defined.

*Added Value of This Study:* Using a trial-like population from a natural history study, in which clinical trial-grade phenotypic data and multi-modal biomarker data were collected, we show that a subset of clinical factors, encapsulated by the ENCALS predictive model score, and serum neurofilament light chain (NfL) are the most powerful prognostic markers when considering either ALSFRS-R functional decline or permanent assisted ventilation (PAV)/tracheostomy-free survival. Importantly, serum NfL adds prognostic value even after adjusting for the ENCALS score, yielding an additional sample size saving of ∼27% in a hypothetical future clinical trial. While serum phosphorylated neurofilament heavy chain (pNfH), urinary p75^ECD^, and plasma miR-181ab each holds some prognostic value, when considered together with the ENCALS score and serum NfL, only p75^ECD^ may yield additional but modest sample size saving.

*Implication of All Available Evidence:* Blood NfL is a validated biomarker for multiple contexts-of-use. As a prognostic marker, it should be used together with clinical predictors, such as the ENCALS predictive model score, in all ongoing and future ALS clinical trials. The utility of urinary p75^ECD^ and plasma miR-181ab is less clear. Serum pNfH, as well as serum uric acid, albumin, creatinine, and C-reactive protein (CRP), provide no additional prognostic information.

## Introduction

Clinical trials in the field of amyotrophic lateral sclerosis (ALS) must consider the phenotypic heterogeneity of disease as well as the related challenge that clinically meaningful outcomes, such as the rate of functional decline and survival, are typically insufficiently sensitive to detect therapeutic effect in the early- and mid-phases of drug development. Biofluid biomarkers that are fit-for-purpose, however, may help to meaningfully address this problem.^1^ In patients with clinically manifest ALS (as opposed to the pre-symptomatic population at elevated risk for ALS, which is beyond the scope of this paper), *prognostic biomarkers* might be used in three broad ways to improve the design and analysis of clinical trials. From a study design perspective, they may be used as eligibility criteria to enrich for a population, in which a therapeutic effect might be most apparent, or to stratify randomisation. They may also be used analytically to adjust for phenotypic heterogeneity, thereby reducing the sample size needed to adequately power a trial using clinical outcome measures.^2^ These approaches are not mutually exclusive, and indeed could be combined, depending on the goals of a particular trial. In addition, *response biomarkers* might be used to demonstrate target engagement or pharmacodynamic effect, and perhaps even serve as surrogates that are reasonably likely to predict a future clinical benefit.

There remains, however, a significant gap between biomarker discovery, analytic validation, and preliminary reports of biomarker performance in samples of convenience on the one hand, and clinical validation on the other hand. The latter entails demonstrating the utility of a biomarker for a well-defined context-of-use in a large, carefully phenotyped clinical cohort.

Prior studies have identified clinical parameters predictive of disease progression ^2^ or survival.^2,3^ Moreover, among patients with ALS, neurofilament light chain (NfL) has emerged as the lead prognostic and response biomarker ^4–8^ for a number of reasons: NfL can reliably be measured in blood, there is a high correlation between blood and cerebrospinal fluid (CSF) concentrations, and empiric data support these contexts-of-use based on results from serum or plasma. There is, however, also persistent interest in the potential prognostic value of other biomarkers, including blood phosphorylated neurofilament heavy chain (pNfH);^9^ urinary p75 neurotrophin receptor extracellular domain (p75^ECD^);^10^ microRNA-181 (miR-181);^11^ and an array of analytes, such as uric acid,^12–20^ albumin,^16^ creatinine,^16,20,21^ and C-reactive protein (CRP),^8,22,23^ that are routinely quantified in the clinical arena.

In this study, we sought to clinically validate the utility of putative prognostic biofluid biomarkers in the context of established clinical prognostic factors. The rationale is that a prognostic biomarker would only be worth quantifying if it adds value to what can be learned from known and readily available clinical parameters. Furthermore, a head-to-head comparison of clinical markers and molecular biomarkers revealed their relative contributions to clinical trial design, analysis, and result interpretation. Finally, we characterised the longitudinal trajectories of a subset of biomarkers, to inform their potential future use as response biomarkers. While prognostic clinical measures and biomarkers may have value in the clinical arena, such individual use of these markers is beyond the scope of the current investigation which is focused on the clinical trial utility of these markers,

## Methods

### Study Population

Patients with ALS were enroled (between 2014 and 2019) at 12 centers in the United States and 1 center in South Africa through the prospective Phenotype-Genotype-Biomarker (PGB) study (registered at clinicaltrials.gov NCT02327845) of the Clinical Research in ALS and Related Disorders for Therapeutic Development (CReATe) Consortium. The PGB study enrolled 705 patients with ALS (n=472), primary lateral sclerosis (n=47), progressive muscular atrophy (n=20), hereditary spastic paraplegia (n=162) and other related disorders (n=4). The goal was to evaluate participants serially over a period of 1.5-2 years to acquire longitudinal phenotypic data. Those with ALS, ALS-FTD and PMA were to be seen at Baseline, and Months 3, 6, 12 and 18; those with PLS, HSP and multisystem proteinopathy were to be seen at Baseline and Months 6, 12, 18 and 24. Biological samples (blood and urine, as well as cerebrospinal fluid when willing) were collected at all study visits. Periodic medical record reviews, in addition to direct communication with patients, were performed as needed to ascertain the timing of survival endpoint (permanent assisted ventilation [PAV; non-invasive ventilation > 23 hours/day], tracheostomy, or death).

While PGB was designed to be broadly inclusive, the subset of patients with ALS who met common trial eligibility criteria were designated as “trial-like” and served as the basis for this report. Key inclusion/exclusion criteria included: diagnosis of ALS according to El Escorial criteria, permitting those with cognitive or behavioural impairment (ALSci or ALSbi, respectively), but excluding ALS-FTD; less than 3 years from onset of weakness; and an erect slow vital capacity (SVC) of ≥50% predicted. All patients with ALS in PGB who met these criteria were included in the current report.

### Clinical Assessments

The ALS Functional Rating Scale-Revised (ALSFRS-R), a 48-point scale that includes bulbar, gross motor, fine motor, and respiratory domains,^24^ was the principal measure of functional status. Symptom onset was defined based on the first appearance of weakness or impaired motor function. The estimated rate of change in ALSFRS-R between symptom onset and baseline (ΔFRS), was defined as (48-baseline ALSFRS-R/months since symptom onset).^25^ Respiratory muscle function was quantified with slow vital capacity (SVC) in the erect position. Alternate versions of the North American version of the Edinburgh Cognitive and Behavioural ALS Screen (ECAS), including informant report, was used to evaluate cognitive and behavioural function.^26^ All evaluators were trained and certified for the performance of each of these outcome measures. Biological sex, as well as race and ethnicity, were self-reported.

### Ethics

The University of Miami institutional review board (IRB), which served as the central IRB for CReATe, approved the study for all US sites study (protocol # 20160603) and the University of Cape Town Health Sciences Human Research Ethics Committee approved the study in South Africa (REF number 165/2017). All participants provided written informed consent.

### Biological Samples

Biological specimens were collected, processed, and stored according to strict standard operating procedures. Briefly, blood was collected in serum-separating BD vacutainers and allowed to clot upright at room temperature for 1–2 hours. Following centrifugation (1,750 g for 10 min at 4°C), serum was aliquoted into cryogenic sterile freestanding conical microtubes (Nalgene or Bio Plas Inc.) and stored at −80°C. Plasma was collected in K2 EDTA tubes, centrifuged at 1,750g for 10 minutes at 4°C within 2 hours of collection, and aliquoted for storage at −80°C. Urine was collected in a sterile collection cup, gently swirled, and transferred to cryovials for immediate storage at −80°C.

### Biomarker and Genetic Assays

Serum NfL and pNfH concentrations were quantified using the Simoa NfL and pNfH assays in the laboratory of an author (AM). Established protocols for NfL (Simoa Nf-L Advantage Kit-102258, Quanterix) and pNfH (Simoa pNF-Heavy Discovery Kit - 102669) analysis were used. Each plate contained calibrators and quality controls. Samples were diluted to fall within the range of the standard curve.

Urinary p75^ECD^ was quantified by ELISA in the laboratory of an author (MLR) as previously described.^10^ Briefly, urinary p75^ECD^ was measured by a sandwich ELISA, that used a capture monoclonal antibody (MLR1 at 8mg/ml) made to the extracellular region of p75 ^27^ in Carbonate-Carbonate coating buffer (Ph 9.6). Another monoclonal antibody (NGFR5) to p75 ^28^ was used as the detection antibody and biotinylated as per the manufacturer’s instructions (Thermo Fisher Scientific Australia, #UG283022) and used at 2.0mg/ml in the assay. Human p75^ECD^ standard was from R&D Systems (Lys29-Asn250; #367-NR). BlockAce (BioRad, BUF-029) was used as blocking and sample buffer. The enzyme reaction was achieved using streptavidin horseradish peroxidase (Jackson ImmunoResearch Laboratories, #JIO16030084) diluted to 1.0 mg /ml and colour developed using tetramethylbenzidine (A:B; BioRad Australia, #1721067). The entire ELISA was accomplished as previously described ^29^ on a Hamilton Starlet Robot, integrated with a Biotek 405 washer, and an MD reader (450nm); two calibrator human urine samples with known p75^ECD^ levels were included on each plate, and if the results from either had greater than 20% coefficient of variation, the results from the plate were rejected. The results were reported as ng p75^ECD^/ ml urine and corrected by creatinine (mg/ml; measured by calorimetric method using Enzo Life Sciences Creatinine Kits (ADI-907-030A) as per the manufacturer’s instructions). Samples with urinary creatinine below 0.3 ± 0.03 mg/ml or above 3.0 ± 0.3 mg ml were rejected as per World Health Organization guidelines ^30^. Final results are reported as ng p75^ECD^ /mg creatinine.

Total RNA was extracted from plasma using the miRNeasy Micro Kit (Qiagen cat. 217084) and quantified with a Qubit fluorometer using the RNA Broad Range Assay Kit (Thermo Fisher Scientific cat. Q10210). For small RNA next-generation sequencing, libraries were prepared from 7.5Ong of total RNA using the QIAseq miRNA Library Kit (cat. 331505) and QIAseq miRNA NGS 48 Index IL (Qiagen cat. 331592) by an experimenter who was blinded to the identity of samples. Precise linear quantification of miRNA gained by UMIs of random 12 nucleotides after 3′ and 5′ adapter ligation, within the reverse transcription primers. cDNA libraries were amplified by PCR for 22 cycles, with a 3′ primer that includes a six-nucleotide unique index, followed by on-bead size selection and cleaning. Library concentration was determined with a Qubit fluorometer (dsDNA High Sensitivity Assay Kit, Thermo Fisher Scientific, cat. Q32851) and library size with TapeStation D1000 (Agilent, cat. Catalog number: Q32851). Libraries with different indices were multiplexed and sequenced on a NovaSeq SP100 (Illumina), with 75-bp single read and 6-bp index read. Human miRNA sequences were mapped using GeneGlobe (Qiagen), normalized with the DESeq2 package and corrected for the library preparation batch. Plasma miR-181a and miR-181b were quantified by small RNA next-generation sequencing in the laboratory of an author (EH) as previously described,^11^ and summarised as miR-181ab, the combined expression of miR-181a and miR-181b. Serum uric acid, albumin, creatinine, and CRP were assayed using the Roche Cobas C Analyzer in the Clinical Chemistry Laboratory at the University of Miami. All biomarker studies were performed blind to clinical outcomes.

The presence of a *C9orf72* repeat expansion was determined in the laboratory of an author (RR) using a two-step protocol, including a fluorescent PCR fragment-length analysis and a repeat-primed PCR, with previously described oligos (ThermoFisher), as described elsewhere.^31^ The PCR reactions (Qiagen) for both assays included Betaine and DMSO additives (MilliporeSigma). The FAM labeled products were run on a 3730xl DNA Analyzer (Applied Biosystems) and sized with Genescan 400 using Genemapper software (ThermoFisher).

### Statistics

Longitudinal change in ALSFRS-R total score, serum NfL, serum pNfH, and urinary p75^ECD^ were estimated in unadjusted mixed model repeated-measures analyses with visits windowed to the closest planned assessment time (at 3, 6, 12, and 18 months) and in unadjusted mixed model random-slopes analyses using the observed assessment times. The repeated-measures model included a fixed effect of visit and assumed unstructured person-level variance-covariance among repeated observations. The random-slopes model included a fixed effect of time and assumed unstructured variance-covariance for the person-level random intercepts and slopes. Biomarker concentrations were log-transformed prior to analysis and estimates were back-transformed. Back-transformation of visit-specific estimates yield values on the original scale of measurement. Back-transformation of slopes or changes from baseline yield geometric mean ratios which were further transformed by subtracting 1 and multiplying by 100 to express as deviations in percentage change from 100%.

We examined an array of clinical measures (sex, onset age, bulbar onset, diagnostic delay, ΔFRS [estimated rate of change in ALSFRS-R between symptom onset and baseline],^25^ baseline age, ALSFRS-R total score, slow vital capacity, ECAS-derived scores, and ENCALS predictor score) and biofluid biomarkers (serum NfL, serum pNfH, urinary p75^ECD^, serum uric acid, serum albumin, serum creatinine, serum CRP, plasma miR-181ab) as potential prognostics of rate of disease progression as measured by ALSFRS-R total score and of PAV/tracheostomy-free survival. We derived five scores from baseline ECAS assessments: total score, ALS-specific score, ALS non-specific score, and dichotomous designations of cognitive impairment (ALSci) and behavioural impairment (ALSbi) defined according to the revised Strong criteria ^32^ and implemented in the PGB study.^33^ ALSci and ALSbi designations were restricted to English-speaking participants for whom robust normative data permitted reliable designation.^26^ The ENCALS linear predictive model score ^3^ (hereinafter “ENCALS predictor score” or “ENCALS score”) combines information from 8 clinical variables (ΔFRS, bulbar onset, diagnostic delay [months from symptom onset to diagnosis], age at onset, El Escorial definite ALS, presence of FTD, presence of a *C9orf72* repeat expansion, and percent predicted vital capacity). Plasma miR-181ab was evaluated as a continuous measure, split at the median (24,590 UMI in the current study), and as defined by Magen et al ^11^ where miR181-ab was defined as a poor prognostic when above the threshold of 39,300 UMI among those in the middle tertile of NfL concentration (NfL 59-109.8pg/ml) ^11^, and as defined by Magen et al but using the median miR181ab value and the middle NfL tertile (44.8-80.8pg/ml) from the current study.

Prognostic markers were assessed for their ability to predict the rate of progression in ALSFRS-R total score in random-slopes analyses and to predict PAV/tracheostomy-free survival by Kaplan-Meier product-limit estimates and by Cox proportional hazards regression. In survival analyses, time at risk began at the baseline visit (time zero) and continued to time last known alive or time of PAV, tracheostomy, or death, if observed. Each model included one prognostic. Continuous prognostics were evaluated both as continuous predictors after standardizing to unit variance and when divided into quartiles. We focused on analyses after dividing prognostics into quartiles (or fewer levels – e.g., for binary measures, where only 2 levels are possible) to avoid the strong assumption of a linear association with rate of progression and survival across the full range of a given prognostic and to permit comparison of all prognostics in a common framework. Models were either unadjusted, adjusted for established core clinical predictors (bulbar onset, ΔFRS, and diagnostic delay for functional decline; plus baseline age for survival), adjusted for ENCALS predictor score, or adjusted also for serum NfL. The adjusted models sharpened estimates by accounting for known sources of variation and addressed whether a given prognostic provided new information independent of known predictors of progression and survival. Wald confidence intervals were used for estimates from random-slopes models. Complementary log-log confidence intervals were used for estimates of median survival time. Profile likelihood confidence intervals were used for estimates of hazard ratios.

In addition to estimating the clinical utility of each potential prognostic biomarker, we quantified the proportional sample size saving that would result from adjusting for a given biomarker in a hypothetical clinical trial. Reductions in sample size requirements based on a normal approximation for a hypothetical clinical trial testing for slowing of ALSFRS-R progression, analyzed in a random slopes model, were estimated based on reductions in standard error estimates of the estimated slopes and resulting increases in the effect size after inclusion of a given prognostic marker as a linear predictor. The proportional savings in sample size assume a consistent but arbitrary allocation ratio, type 1 error control, and power between designs and an assessment schedule similar to the present cohort. For any given choice of allocation ratio, type 1 error control, and power, the relative sample size required for two trials with equivalent assessment schedule differs only as a function of the ratio of the respective effect sizes for tests of the primary outcome, in the present case the estimated slope of ALSFRS-R. Note that effect size ratios rather than variance ratios were used due to small variation in estimated slopes when adding covariates.

A post-hoc analysis of the association between serum NfL and rate of progression in ALSFRS-R total score was performed using a cubic smoothing spline through the empirical Bayes ALSFRS-R slope estimates from an unadjusted random-slopes analysis and using a partial-linear spline in a longitudinal random-slopes analysis. Knots for the partial-linear spline were chosen post-hoc, based on visual inspection, at 40 and 100 pg/mL to approximate the shape of the cubic smoothing spline.

Analyses were performed using SAS (version 9.4, SAS Institute, Cary NC) and R (version 4.0.3, R Foundation for Statistical Computing, Vienna, Austria). Comparison-wise p-values are reported with nominal significance at two-tailed p < 0.05. Results significant after correction by Holm-Bonferroni stepdown adjustment for multiple comparisons over 28 prognostic markers are indicated.

### Role of Funders

The funders of the study had no role in study design, data collection, statistical analysis, results interpretation, or writing of the report.

## Results

### Study Population

A total of 203 patients with ALS were included, with a mean (±SD) age of 57.1 (±12) years and a slight male preponderance (55%). A genetic cause of ALS was identified in 24 (12%), most commonly a *C9orf72* hexanucleotide repeat expansion (n=20; 10%). Median disease duration (time since symptom onset) at baseline was 14.4 months, with a mean SVC of 85% (±17) predicted (Table 1a). ALSFRS-R declined by an average (±SE) of 0.89 (±0.05) points/month (Figure 1a) with median (Q1-Q3) follow-up of 10.1 (5.8-16.3) months. SVC declined by an average (±SE) of 1.8% (±0.15) predicted per month. 93 (46%) patients reached a survival endpoint (PAV, tracheostomy, or death), with a median (Q1-Q3) survival time of 30.1 (17.4-47.7) months observed from follow-up of 17.4 (10.6-29.9) months. 110 patients were censored (25 study completion, 15 loss to follow-up, 11 withdrawal/dropout, and 59 administrative study closure).

**Figure 1.**
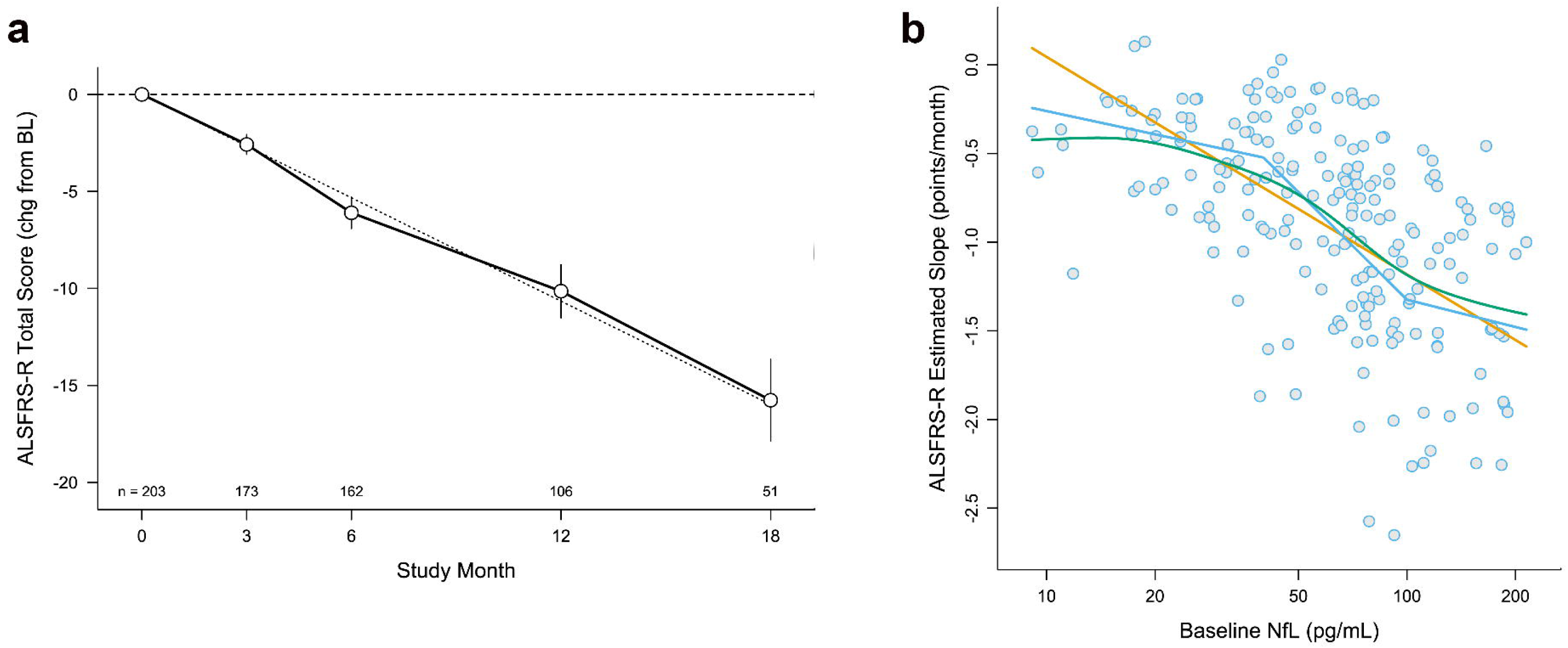
ALSFRS-R Slope and its Relationship to Baseline NfL **(a)** Random slopes model of ALSFRS-R over time, with errors bars showing 95% confidence intervals (CI). Faint grey dotted line illustrates the linear estimate of change in ALSFRS-R over time. **(b)** Relationship between baseline serum NfL (measured in duplicate) and future rate of progression of the ALSFRS-R (Spearman correlation coefficient = −0.57, 95% CI −0.66 to −0.47, p<0.0001) among n=203 study participants. The straight orange line shows the linear prediction. The bent blue line represents a partial-linear spline with knots chosen post-hoc at 40 and 100 pg/mL. The smooth green curve is a smoothing spline through the empirical Bayes slope estimates.

**Table 1a.** Baseline demographic and key clinical features.

| Characteristic |  | Total N=203 |
| --- | --- | --- |
| Sex | Male | 112 (55.2%) |
|  | Female | 91 (44.8%) |
| Race | White | 179 (88.2%) |
|  | Black | 15 (7.4%) |
|  | Asian | 4 (2.0%) |
|  | Other | 5 (2.5%) |
| Ethnicity | Hispanic/Latino | 36 (17.7%) |
|  | Non-Hispanic/Latino | 167 (82.3%) |
| Gene with a pathogenic variant | *C9orf72* | 20 (9.9%) |
|  | *SOD1* | 3 (1.5%) |
|  | *TARDBP* | 1 (0.5%) |
|  | [Unknown]<sup>2</sup> | 179 (88.2%) |
| Age at onset, years | Mean ± SD (Range) | 57.1 ± 12.0 (15-82) |
| Bulbar symptoms at onset | No | 147 (72.4%) |
|  | Yes | 56 (27.6%) |
| Diagnostic delay, months | Median (Q1-Q3) | 8.2 (5.2-13.8) |
| Time from symptom onset to baseline, months | Median (Q1-Q3) | 14.4 (10.5-22.5) |
| Baseline ΔFRS, points/month | Median (Q1-Q3) | 0.60 (0.4-0.9) |
| Baseline age, years | Mean ± SD (Range) | 58.5 ± 12.1 (17-83) |
| Baseline El Escorial category | Clinically definite ALS | 45 (22.2%) |
|  | Clinically probable ALS | 95 (46.8%) |
|  | Other | 63 (31.0%) |
| Baseline ALSFRS-R total score | Mean ± SD (Range) | 37.9 ± 5.7 (15-48) |
| Baseline SVC, %predicted | Mean ± SD (Range) | 85.3 ± 17.2 (52.0-135.5) |
| Baseline ECAS total score<sup>1</sup> | Mean ± SD (Range) | 110.2 ± 12.4 (44-130) |
| Baseline ECAS ALS-specific score<sup>1</sup> | Mean ± SD (Range) | 81.8 ± 10.3 (35-97) |
| Baseline ECAS ALS non-specific score<sup>1</sup> | Mean ± SD (Range) | 28.4 ± 3.7 (9-35) |
| Baseline cognitive impairment, by ECAS | No | 137 (87.8%) |
|  | Yes | 19 (12.2%) |
|  | [n/a]<sup>3</sup> | 47 (--) |
| Baseline behavioural impairment, by ECAS | No | 95 (84.8%) |
|  | Yes | 17 (15.2%) |
|  | [n/a]<sup>4</sup> | 91 (--) |
| Survival duration from baseline, months | Median (Q1-Q3) | 30.1 (17.4-47.7) |
| PAV, tracheostomy, or death occurrence | No | 110 (54.2%) |
|  | Yes | 93 (45.8%) |
| Number of sample collections | 2 | 48 (23.6%) |
|  | 3 | 56 (27.6%) |
|  | 4 | 47 (23.2%) |
|  | 5 | 52 (25.6%) |

<sup>1</sup> Among English speakers (n=171)
<sup>2</sup> No pathogenic variant identified (by *C9orf72* testing and whole genome sequencing) in known disease-causing genes.
<sup>3</sup> Not available, because a non-English version of ECAS was completed or if there was insufficient information to determine impairment status.
<sup>4</sup> Not available, because a non-English version of ECAS was completed, caregiver did not complete ECAS, or if there was insufficient information to determine impairment status.
SD = Standard deviation. Q1 = 1<sup>st</sup> quartile. Q3 = 3<sup>rd</sup> quartile. ECAS = Edinburgh Cognitive and Behavioural ALS Screen. SVC = Slow vital capacity. PAV = Permanent assisted ventilation.

**Table 1b.** Baseline biomarker data.

| **Biomarker** | **N** | **Mean ±SD** | **Median (Q1, Q3)** | **Range** |
| --- | --- | --- | --- | --- |
| Serum NfL (pg/mL) | 203 | 73.9 ± 47.0 | 67.9 (37.9, 92.5) | 9.1 – 214 |
| Serum pNfH (pg/mL) | 203 | 598 ± 718 | 267 (110, 924) | 3.4 – 4,177 |
| Serum NfL/pNfH ratio | 203 | 0.52 ± 1.56 | 0.18 (0.09, 0.50) | 0.020 – 20.9 |
| Urinary p75<sup>ECD</sup> (ng/mg creatinine) | 160<sup>1</sup> | 5.54 ± 2.42 | 5.05 (3.93, 6.44) | 1.53 – 16.1 |
| Serum uric acid (mg/dL) | 203 | 5.19 ± 1.31 | 5.00 (4.20, 6.20) | 2.60 – 8.90 |
| Serum creatinine (mg/dL) | 203 | 0.78 ± 0.20 | 0.77 (0.66, 0.90) | 0.29 – 1.59 |
| Serum albumin (g/dL) | 203 | 4.55 ± 0.34 | 4.50 (4.30, 4.80) | 3.60 – 5.80 |
| Serum CRP (mg/dL) | 203 | 0.27 ± 0.35 | 0.10 (0.10, 0.50) | 0.10 – 2.30 |
| Plasma miR-181a (UMI) | 201 | 451 ± 208 | 418 (312, 552) | 124 – 1,699 |
| Plasma miR-181b (UMI) | 201 | 66.2 ± 31.4 | 61.2 (44.8, 79.7) | 13.8 – 263 |
| Plasma miR-181ab (UMI<sup>2</sup>) | 201 | 34,663 ± 39,372 | 24,590 (13,638, 42,836) | 3,148 – 447,480 |

UMI = unique molecular identifier
<sup>1</sup> Urinary p75<sup>ECD</sup> only available from a subset of study participants.

### Biomarker Profiles

Baseline serum NfL concentrations ranged from 9 to 214 pg/mL (Table 1b) and correlated with subsequent rates of ALSFRS-R decline (Spearman r=-0.57, 95% CI [−0.66, −0.47], p<0.0001, Figure 1b). Over the course of follow-up, serum NfL increased by an average (95% CI) of 0.98% [0.57%, 1.38%] per month (Table 2, Figure 2a). Baseline serum pNfH concentrations ranged from 3.4 to 4,177 pg/mL (Table 1b) and increased by an average of 0.45% [−0.12%, 1.03%] per month (Table 2, Figure 2b). Baseline urinary p75^ECD^ levels ranged from 1.5 to 16.2 ng/mg creatinine (Table 1b) and increased by an average of 2.59% [2.01%, 3.17%] per month (Table 2, Figure 2c). Baseline plasma miR-181ab (product of miR-181a and miR-181b) concentration ranged from 2,875 to 431,004 unique molecular identifiers (UMIs) (Table 1b). Baseline concentrations of serum uric acid, albumin, creatinine, and CRP are summarised in Table 1b.

**Figure 2.**
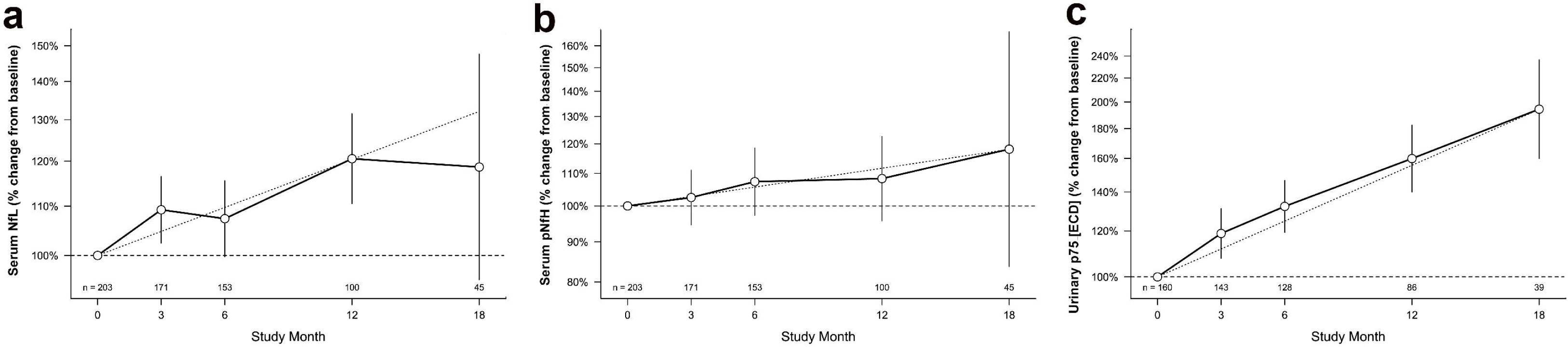
Longitudinal Biomarker Trajectories Longitudinal trajectories of **(a)** serum NfL; **(b)** serum pNfH; and **(c)** urinary p75^ECD^. Y-axis shows percent change in each biomarker compared to baseline, plotted on a log scale. The faint grey dotted line illustrates the linear estimate of biomarker change over time. Error bars represent 95% confidence intervals (CI), widened at later time points due to participant attrition over time and fewer biomarker data available. NfL and pNfH were measured in duplicate; p75^ECD^ quantified with a median of 3 replicates. Single NfL and pNfH values from the 18-month visit of a single participant have been excluded (see footnote to Table 2 for detailed explanation).

**Table 2.** Longitudinal biomarker trajectories.

| **Biomarker** | **Increase per month, relative to baseline** |  |  |
| --- | --- | --- | --- |
|  | **Mean** | **(95% CI)** | **P-value** |
| Serum NfL (pg/mL)<sup>1</sup> | 0.98% | (0.57%, 1.38%) | <0.0001 |
| Serum pNfH (pg/mL)<sup>1</sup> | 0.45% | (-0.12%, 1.03%) | 0.12 |
| Serum NfL/pNfH ratio | 0.44% | (-0.09%, 0.97%) | 0.10 |
| Urinary p75<sup>ECD</sup> (ng/mg creatinine) | 2.59% | (2.01%, 3.17%) | <0.0001 |

Values are unadjusted for core clinical covariates
<sup>1</sup> One substantial outlier, from the 18-month visit (i.e. visit 5) of a participant, was excluded.

Baseline biomarker results, stratified by *C9orf72* status and by sex are summarised in eTable 1 and eTable 2 respectively, with longitudinal changes in biomarkers stratified by sex in eTable 3. Correlations among all prognostics at baseline are summarised in eTable 4.

### Prognostic Markers for Survival

In univariate models, the strongest predictors of survival were the ENCALS score, baseline serum NfL, and ΔFRS. Median survival among those with ENCALS predictor scores in the lowest vs. highest quartiles (i.e., lowest vs. highest predicted risk of PAV/tracheostomy-free survival) were 48 vs. 17 months (Table 3, Figure 3a). Median survival among those with ΔFRS slopes in the lowest vs. highest quartiles (i.e., slowest vs. fastest pre-baseline slope) were 47 vs. 17 months (Table 3, Figure 3b). Median survival among those with baseline NfL concentrations in the lowest vs. highest quartiles were 49 vs. 17 months (Table 3, Figure 3c). Median survival among those with baseline miR181ab above vs. below 24,590 UMI were. 23 vs. 35 months (Table 3, Figure 3d). Bulbar onset, baseline ALSFRS-R, and baseline SVC %predicted also predicted survival (Table 3).

**Figure 3.**
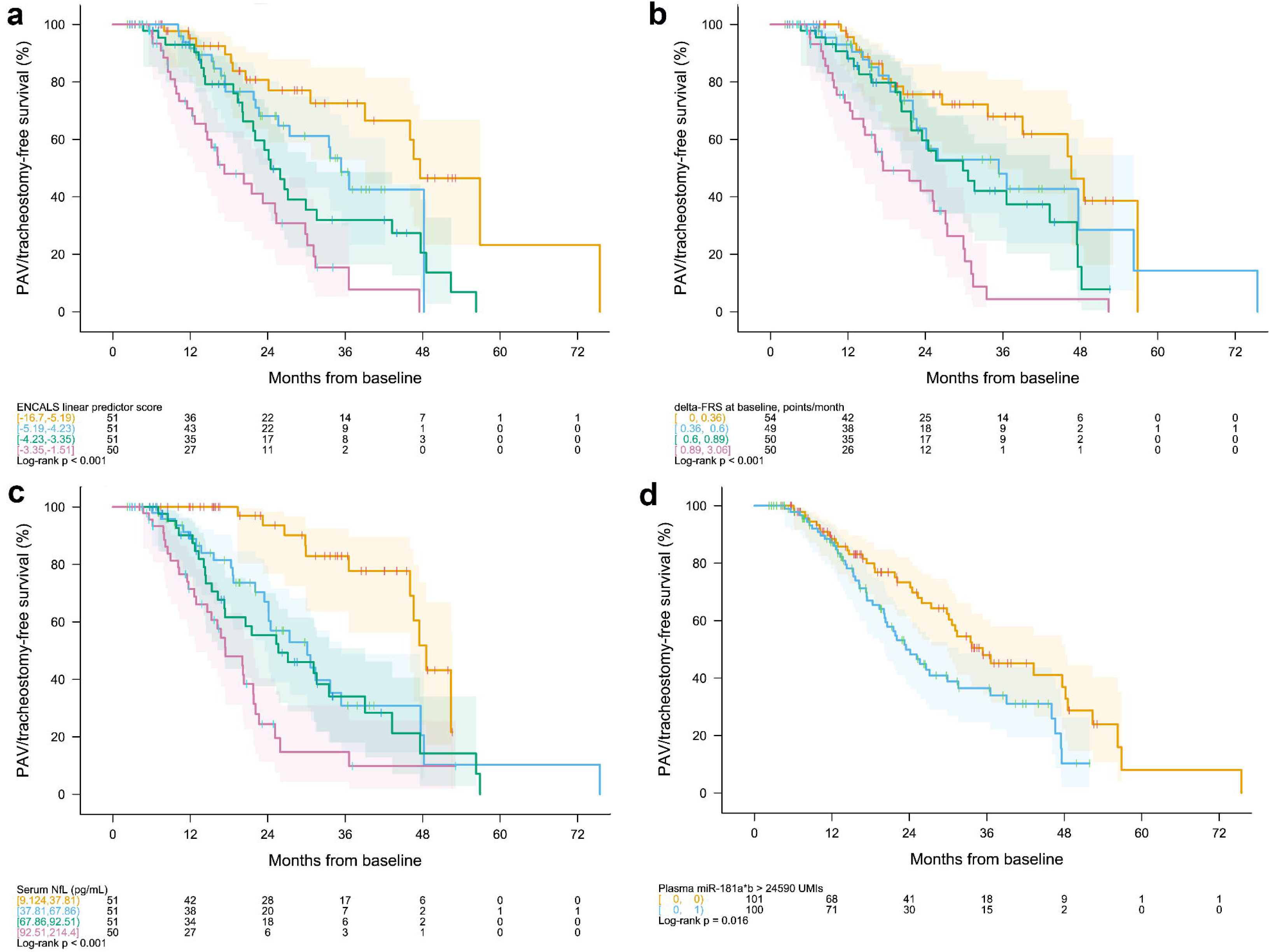
Kaplan-Meier Survival Curves Permanent assisted ventilation (PAV)- and tracheostomy-free survival for **(a)** the ENCALS predictor score, divided into quartiles; **(b)** ΔFRS, divided into quartiles; **(c)** baseline serum NfL, divided into quartiles; and **(d)** baseline plasma miR-181ab dichotomised at the median value of 24,590 UMI. The range of values for each clinical or biological marker within a defined quartile, as well as the number of observations at each time point, are shown below each KM plot. Shading represents pointwise log-log confidence intervals.

**Table 3.**
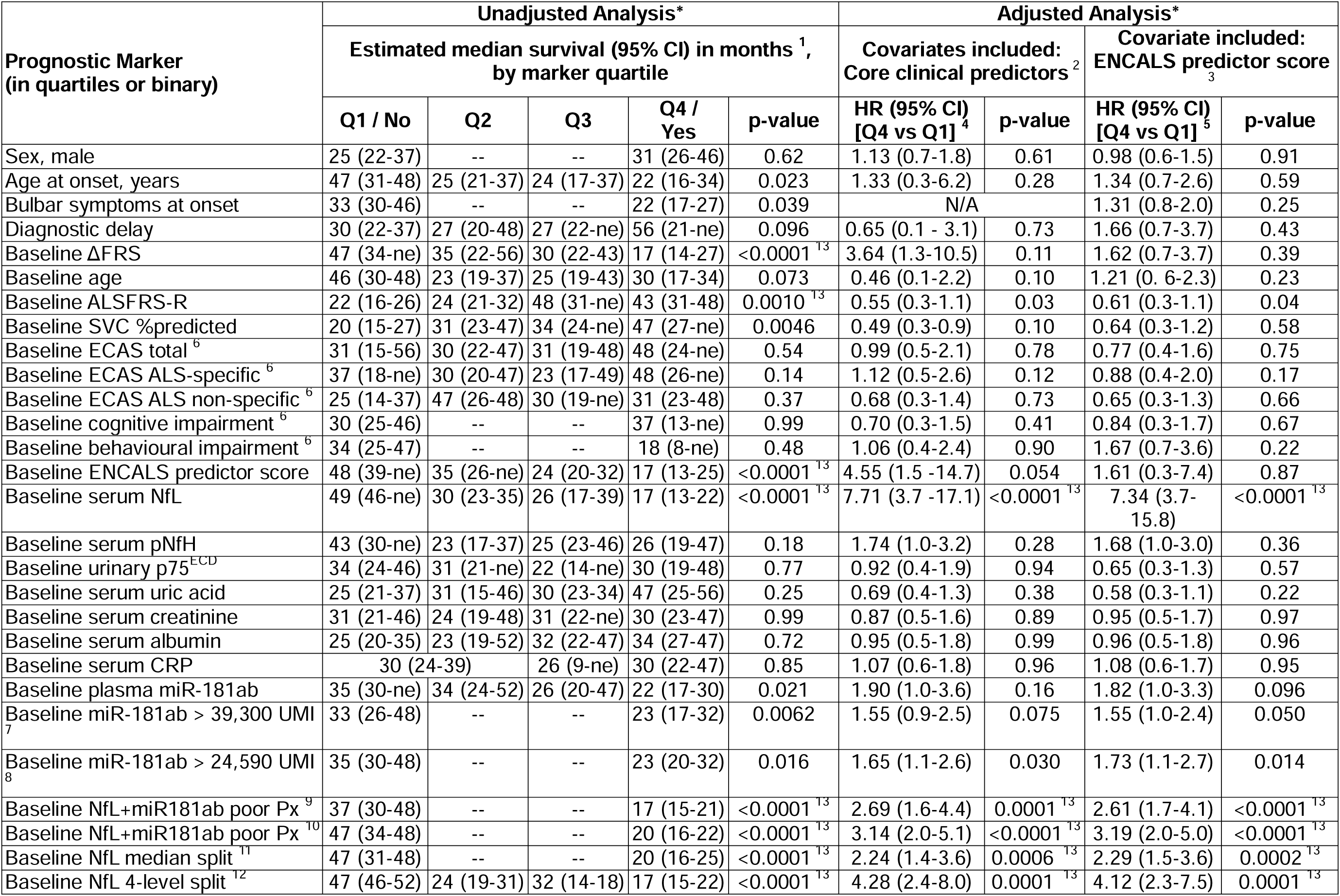
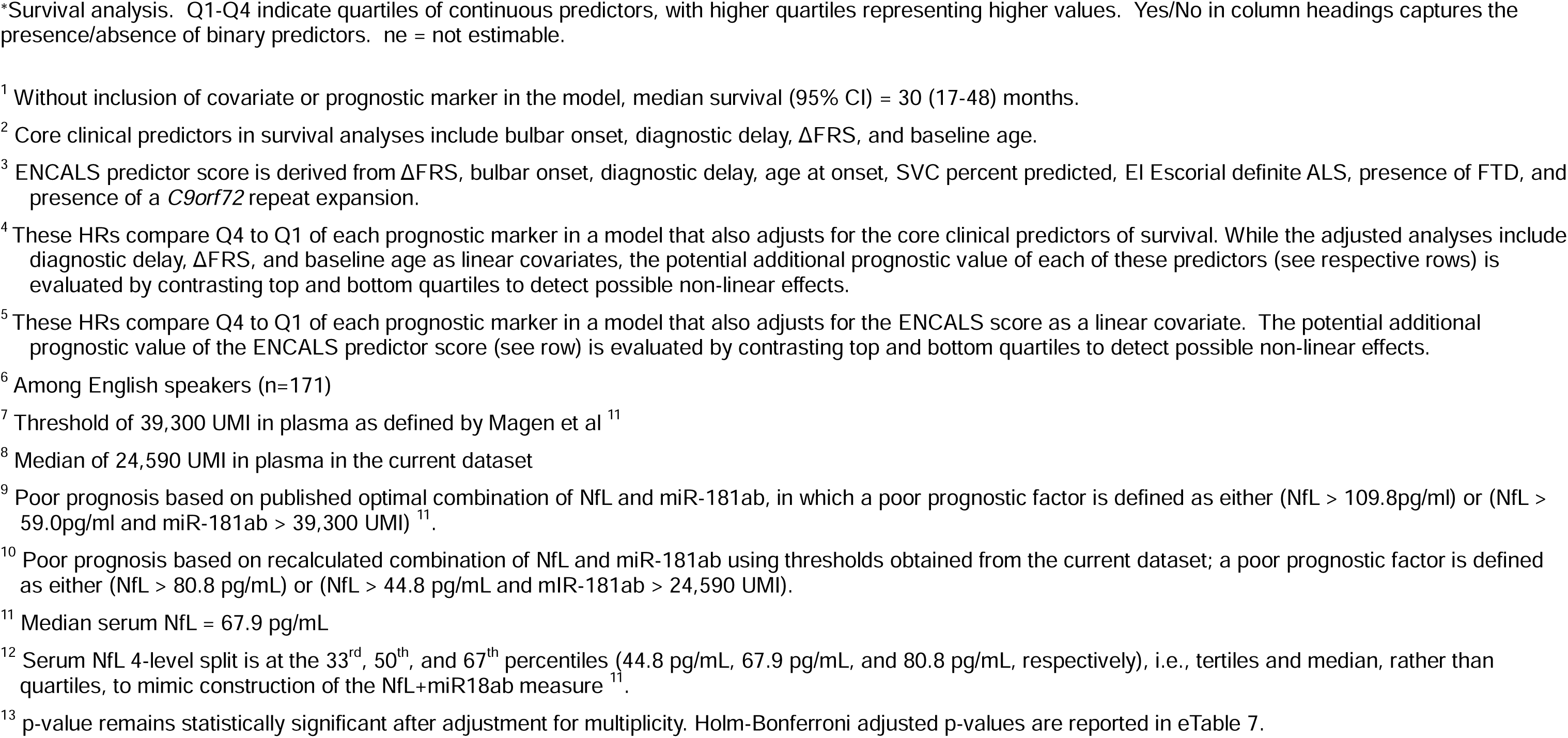
Prognostic markers of survival.

| Prognostic Marker<br>(in quartiles or binary) | Unadjusted Analysis* |  |  |  |  | Adjusted Analysis* |  |  |  |
| --- | --- | --- | --- | --- | --- | --- | --- | --- | --- |
|  | Estimated median survival (95% CI) in months <sup>1</sup>,<br>by marker quartile |  |  |  |  | Covariates included:<br>Core clinical predictors <sup>2</sup> |  | Covariate included:<br>ENCALS predictor score <sup>3</sup> |  |
|  | Q1 / No | Q2 | Q3 | Q4 /<br>Yes | p-value | HR (95% CI)<br>[Q4 vs Q1] <sup>4</sup> | p-value | HR (95% CI)<br>[Q4 vs Q1] <sup>5</sup> | p-value |
| Sex, male | 25 (22-37) | -- | -- | 31 (26-46) | 0.62 | 1.13 (0.7-1.8) | 0.61 | 0.98 (0.6-1.5) | 0.91 |
| Age at onset, years | 47 (31-48) | 25 (21-37) | 24 (17-37) | 22 (16-34) | 0.023 | 1.33 (0.3-6.2) | 0.28 | 1.34 (0.7-2.6) | 0.59 |
| Bulbar symptoms at onset | 33 (30-46) | -- | -- | 22 (17-27) | 0.039 | N/A |  | 1.31 (0.8-2.0) | 0.25 |
| Diagnostic delay | 30 (22-37) | 27 (20-48) | 27 (22-ne) | 56 (21-ne) | 0.096 | 0.65 (0.1 - 3.1) | 0.73 | 1.66 (0.7-3.7) | 0.43 |
| Baseline ΔFRS | 47 (34-ne) | 35 (22-56) | 30 (22-43) | 17 (14-27) | <0.0001 <sup>13</sup> | 3.64 (1.3-10.5) | 0.11 | 1.62 (0.7-3.7) | 0.39 |
| Baseline age | 46 (30-48) | 23 (19-37) | 25 (19-43) | 30 (17-34) | 0.073 | 0.46 (0.1-2.2) | 0.10 | 1.21 (0.6-2.3) | 0.23 |
| Baseline ALSFRS-R | 22 (16-26) | 24 (21-32) | 48 (31-ne) | 43 (31-48) | 0.0010 <sup>13</sup> | 0.55 (0.3-1.1) | 0.03 | 0.61 (0.3-1.1) | 0.04 |
| Baseline SVC %predicted | 20 (15-27) | 31 (23-47) | 34 (24-ne) | 47 (27-ne) | 0.0046 | 0.49 (0.3-0.9) | 0.10 | 0.64 (0.3-1.2) | 0.58 |
| Baseline ECAS total <sup>6</sup> | 31 (15-56) | 30 (22-47) | 31 (19-48) | 48 (24-ne) | 0.54 | 0.99 (0.5-2.1) | 0.78 | 0.77 (0.4-1.6) | 0.75 |
| Baseline ECAS ALS-specific <sup>6</sup> | 37 (18-ne) | 30 (20-47) | 23 (17-49) | 48 (26-ne) | 0.14 | 1.12 (0.5-2.6) | 0.12 | 0.88 (0.4-2.0) | 0.17 |
| Baseline ECAS ALS non-specific <sup>6</sup> | 25 (14-37) | 47 (26-48) | 30 (19-ne) | 31 (23-48) | 0.37 | 0.68 (0.3-1.4) | 0.73 | 0.65 (0.3-1.3) | 0.66 |
| Baseline cognitive impairment <sup>6</sup> | 30 (25-46) | -- | -- | 37 (13-ne) | 0.99 | 0.70 (0.3-1.5) | 0.41 | 0.84 (0.3-1.7) | 0.67 |
| Baseline behavioural impairment <sup>6</sup> | 34 (25-47) | -- | -- | 18 (8-ne) | 0.48 | 1.06 (0.4-2.4) | 0.90 | 1.67 (0.7-3.6) | 0.22 |
| Baseline ENCALS predictor score | 48 (39-ne) | 35 (26-ne) | 24 (20-32) | 17 (13-25) | <0.0001 <sup>13</sup> | 4.55 (1.5 -14.7) | 0.054 | 1.61 (0.3-7.4) | 0.87 |
| Baseline serum NfL | 49 (46-ne) | 30 (23-35) | 26 (17-39) | 17 (13-22) | <0.0001 <sup>13</sup> | 7.71 (3.7 -17.1) | <0.0001 <sup>13</sup> | 7.34 (3.7-15.8) | <0.0001 <sup>13</sup> |
| Baseline serum pNfH | 43 (30-ne) | 23 (17-37) | 25 (23-46) | 26 (19-47) | 0.18 | 1.74 (1.0-3.2) | 0.28 | 1.68 (1.0-3.0) | 0.36 |
| Baseline urinary p75<sup>ECD</sup> | 34 (24-46) | 31 (21-ne) | 22 (14-ne) | 30 (19-48) | 0.77 | 0.92 (0.4-1.9) | 0.94 | 0.65 (0.3-1.3) | 0.57 |
| Baseline serum uric acid | 25 (21-37) | 31 (15-46) | 30 (23-34) | 47 (25-56) | 0.25 | 0.69 (0.4-1.3) | 0.38 | 0.58 (0.3-1.1) | 0.22 |
| Baseline serum creatinine | 31 (21-46) | 24 (19-48) | 31 (22-ne) | 30 (23-47) | 0.99 | 0.87 (0.5-1.6) | 0.89 | 0.95 (0.5-1.7) | 0.97 |
| Baseline serum albumin | 25 (20-35) | 23 (19-52) | 32 (22-47) | 34 (27-47) | 0.72 | 0.95 (0.5-1.8) | 0.99 | 0.96 (0.5-1.8) | 0.96 |
| Baseline serum CRP | 30 (24-39) |  | 26 (9-ne) | 30 (22-47) | 0.85 | 1.07 (0.6-1.8) | 0.96 | 1.08 (0.6-1.7) | 0.95 |
| Baseline plasma miR-181ab | 35 (30-ne) | 34 (24-52) | 26 (20-47) | 22 (17-30) | 0.021 | 1.90 (1.0-3.6) | 0.16 | 1.82 (1.0-3.3) | 0.096 |
| Baseline miR-181ab > 39,300 UMI <sup>7</sup> | 33 (26-48) | -- | -- | 23 (17-32) | 0.0062 | 1.55 (0.9-2.5) | 0.075 | 1.55 (1.0-2.4) | 0.050 |
| Baseline miR-181ab > 24,590 UMI <sup>8</sup> | 35 (30-48) | -- | -- | 23 (20-32) | 0.016 | 1.65 (1.1-2.6) | 0.030 | 1.73 (1.1-2.7) | 0.014 |
| Baseline NfL+miR181ab poor Px <sup>9</sup> | 37 (30-48) | -- | -- | 17 (15-21) | <0.0001 <sup>13</sup> | 2.69 (1.6-4.4) | 0.0001 <sup>13</sup> | 2.61 (1.7-4.1) | <0.0001 <sup>13</sup> |
| Baseline NfL+miR181ab poor Px <sup>10</sup> | 47 (34-48) | -- | -- | 20 (16-22) | <0.0001 <sup>13</sup> | 3.14 (2.0-5.1) | <0.0001 <sup>13</sup> | 3.19 (2.0-5.0) | <0.0001 <sup>13</sup> |
| Baseline NfL median split <sup>11</sup> | 47 (31-48) | -- | -- | 20 (16-25) | <0.0001 <sup>13</sup> | 2.24 (1.4-3.6) | 0.0006 <sup>13</sup> | 2.29 (1.5-3.6) | 0.0002 <sup>13</sup> |
| Baseline NfL 4-level split <sup>12</sup> | 47 (46-52) | 24 (19-31) | 32 (14-18) | 17 (15-22) | <0.0001 <sup>13</sup> | 4.28 (2.4-8.0) | 0.0001 <sup>13</sup> | 4.12 (2.3-7.5) | 0.0001 <sup>13</sup> |

\*Survival analysis. Q1-Q4 indicate quartiles of continuous predictors, with higher quartiles representing higher values. Yes/No in column headings captures the presence/absence of binary predictors. ne = not estimable.
<sup>1</sup> Without inclusion of covariate or prognostic marker in the model, median survival (95% CI) = 30 (17-48) months.
<sup>2</sup> Core clinical predictors in survival analyses include bulbar onset, diagnostic delay, ΔFRS, and baseline age.
<sup>3</sup> ENCALS predictor score is derived from ΔFRS, bulbar onset, diagnostic delay, age at onset, SVC percent predicted, El Escorial definite ALS, presence of FTD, and presence of a *C9orf72* repeat expansion.
<sup>4</sup> These HRs compare Q4 to Q1 of each prognostic marker in a model that also adjusts for the core clinical predictors of survival. While the adjusted analyses include diagnostic delay, ΔFRS, and baseline age as linear covariates, the potential additional prognostic value of each of these predictors (see respective rows) is evaluated by contrasting top and bottom quartiles to detect possible non-linear effects.
<sup>5</sup> These HRs compare Q4 to Q1 of each prognostic marker in a model that also adjusts for the ENCALS score as a linear covariate. The potential additional prognostic value of the ENCALS predictor score (see row) is evaluated by contrasting top and bottom quartiles to detect possible non-linear effects.
<sup>6</sup> Among English speakers (n=171)
<sup>7</sup> Threshold of 39,300 UMI in plasma as defined by Magen et al <sup>11</sup>
<sup>8</sup> Median of 24,590 UMI in plasma in the current dataset
<sup>9</sup> Poor prognosis based on published optimal combination of NfL and miR-181ab, in which a poor prognostic factor is defined as either (NfL > 109.8pg/ml) or (NfL > 59.0pg/ml and miR-181ab > 39,300 UMI) <sup>11</sup>.
<sup>10</sup> Poor prognosis based on recalculated combination of NfL and miR-181ab using thresholds obtained from the current dataset; a poor prognostic factor is defined as either (NfL > 80.8 pg/mL) or (NfL > 44.8 pg/mL and miR-181ab > 24,590 UMI).
<sup>11</sup> Median serum NfL = 67.9 pg/mL
<sup>12</sup> Serum NfL 4-level split is at the 33<sup>rd</sup>, 50<sup>th</sup>, and 67<sup>th</sup> percentiles (44.8 pg/mL, 67.9 pg/mL, and 80.8 pg/mL, respectively), i.e., tertiles and median, rather than quartiles, to mimic construction of the NfL+miR18ab measure <sup>11</sup>.
<sup>13</sup> p-value remains statistically significant after adjustment for multiplicity. Holm-Bonferroni adjusted p-values are reported in eTable 7.

In Cox proportional hazards models of time to death or equivalent, we evaluated the prognostic utility of each clinical and biofluid marker when added as quartiles to multivariate models that included either a core set of clinical predictors (bulbar, ΔFRS, and diagnostic delay) or the ENCALS predictor score (Table 3). Results from models with prognostic measures added as linear terms are summarised in eTable 5. When a given prognostic is included both as quartiles and as a linear term among the covariates, the results presented in Table 3 describe any non-linearity in the relationship with survival. Serum NfL remained the strongest predictor. For example, in a model that already includes the ENCALS predictor score, the hazard ratio (HR) for the fourth vs. first quartile values of NfL is 7.3 (Table 3). The addition of NfL as a linear term to an ENCALS-adjusted Cox model, yields a HR of 1.83 for every 1 standard deviation increase in NfL (eTable 5). To examine the prognostic value of plasma miR-181ab in these multivariate models, we considered multiple analytic approaches. The previously published approach, in which a higher miR-181ab is categorised as poor prognostic only for the middle tertile of NfL ^11^, reveals no prognostic value beyond that conferred by NfL alone, whether tertiles from a prior study or the current cohort were used. By contrast, dichotomising at the median value in this cohort (but not the threshold value identified in a prior study) added some prognostic value – with HRs of 1.65 and 1.73, respectively, when miR-181ab was added to the core set of clinical predictors and the ENCALS predictor score (Table 3). None of the other biomarker candidates considered – serum uric acid, albumin, creatinine, or CRP – added prognostic value in survival analyses (Table 3). Similarly, none of our measures of cognitive/behavioural impairment predicted survival (Table 3).

### Prognostic Markers for Functional Decline

In univariate random-slope models of ALSFRS-R decline, ΔFRS, diagnostic delay, ENCALS score, baseline NfL, and baseline pNfH were identified as prognostic markers (Table 4).

**Table 4.**
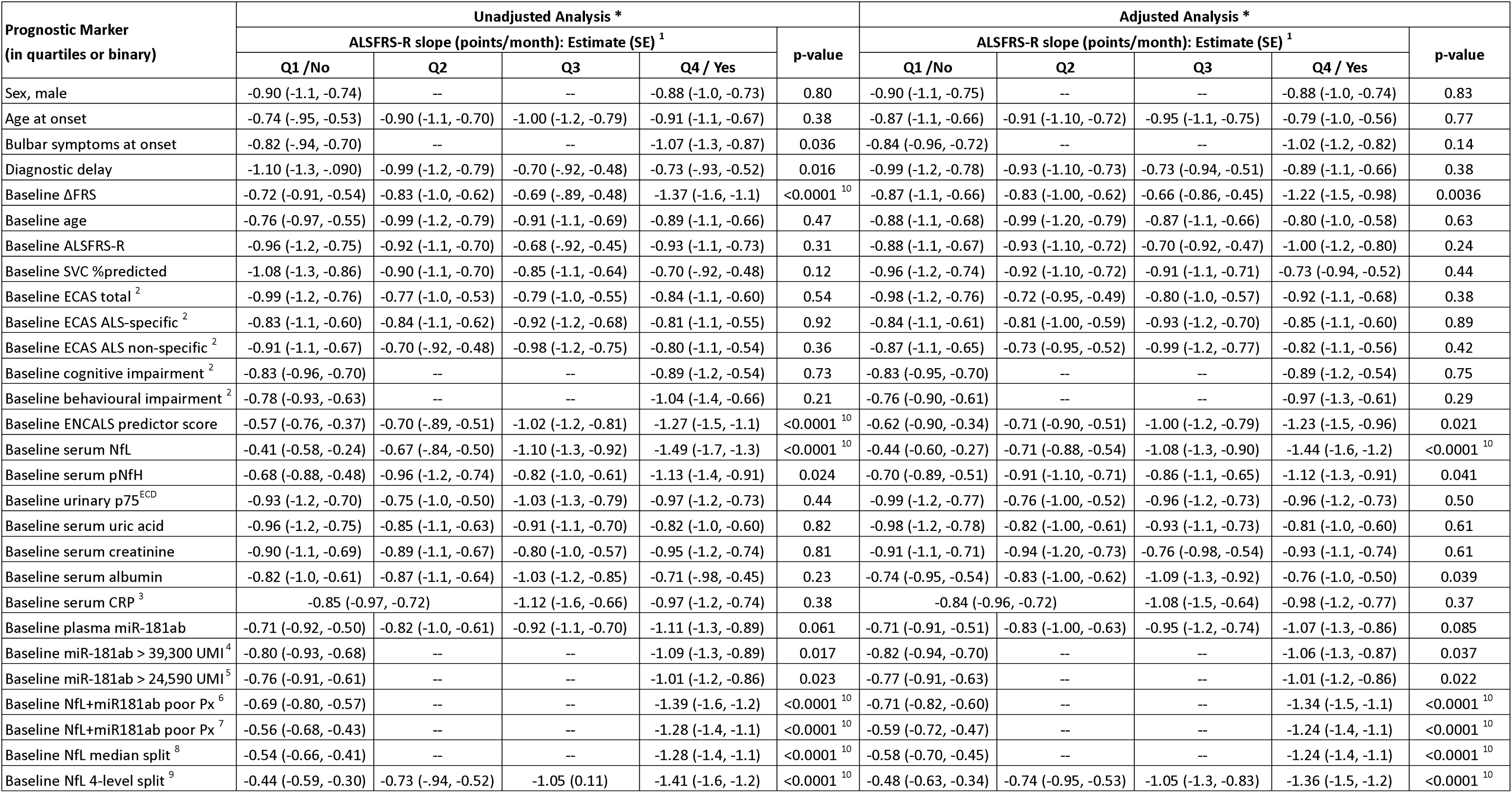
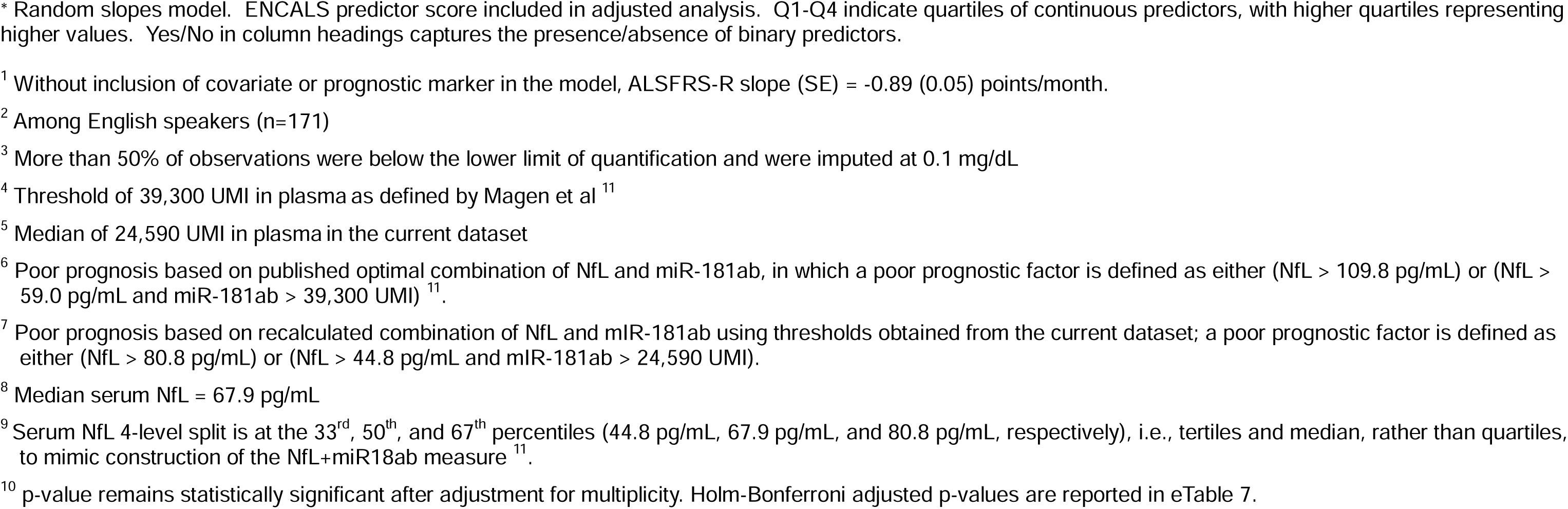
Prognostic markers of functional decline.

Although not developed for predicting functional decline, the ENCALS model predicted differential rates of disease progression that ranged from −0.57 to −1.27 points/month among those with the lowest vs. highest quartile ENCALS scores (Table 4, unadjusted). NfL is also a powerful predictor of future functional decline, with slopes ranging from −0.41 to −1.49 points/month among those with the lowest vs. highest quartiles NfL values (Table 4). Results from models with prognostic measures added as linear terms are summarised in eTable 6.

In multivariate models that already incorporate the ENCALS predictor score, quartiles of baseline serum NfL added substantial prognostic value, with the rate of ALSFRS-R progression ranging from −0.44 to −1.44 points/month among those with the lowest vs. highest quartile values. ΔFRS and serum pNfH added much less prognostic value. Irrespective of the analytic approach, plasma miR-181ab did not add prognostic value beyond that conferred by serum NfL (Table 4). None of the other clinical markers (including measures of cognitive and behavioural impairment), or biomarker candidates considered added prognostic value in random-slopes models of ALSFRS-R functional decline (Table 4).

### Impact of Prognostic Markers on Sample Size Savings for Future Clinical Trials

For the outcome measure of ALSFRS-R slope, the ENCALS model yields a 9% sample size saving, compared to 30.9% for NfL alone (Table 5). The combination of ENCALS and NfL yields a ∼34% saving in sample size. In random slope models of ALSFRS-R that incorporate either the core clinical predictors plus NfL, or ENCALS predictor score plus NfL, the addition of urinary p75^ECD^ yields an additional ∼4% sample size saving, suggesting a modest additional utility of this prognostic marker (with the caveat that this conclusion is based on incomplete baseline data for p75^ECD^ in this sample). The addition of serum pNfH or plasma miR-181ab, however, yielded no additional sample size saving, indicating that in multivariate models that incorporate clinical predictors and NfL, these latter biomarkers add little prognostic value when the ALSFRS-R slope is the outcome measure (Table 5). None of the other clinical measures (including those of cognitive or behavioural impairment) or biomarker candidates yielded sample size savings when considered as prognostic markers.

**Table 5.**
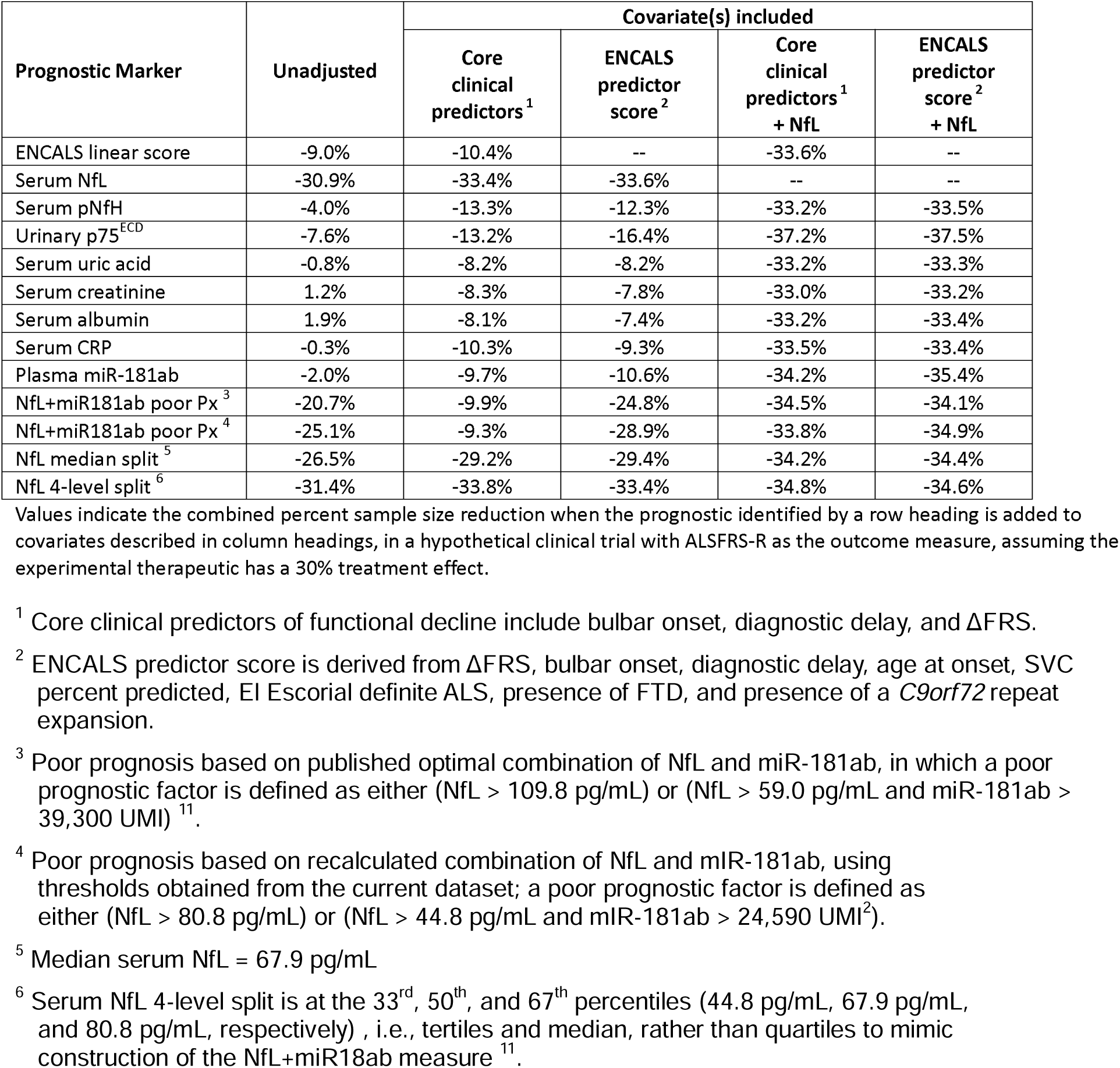
Estimated total sample size savings in a random slopes model of ALSFRS-R progression that includes the prognostic marker and covariate(s)

### A Practical Approach to Incorporating NfL into Trial Design

The relationship between baseline NfL and future rate of functional decline, as measured by the slope of the ALSFRS-R, is not linear (Figure 1b). In this dataset, the sigmoidal relationship yields an estimate of thresholds that might be used either as eligibility criteria (for trial enrichment) or to facilitate stratifying randomisation (to ensure equal balance of NfL-predicted faster and slower disease progression rates across treatment groups). Baseline NfL levels <40 pg/mL corresponded to a future ALSFRS-R slope of ∼0.5 points/month (i.e. slow progression), whereas baseline levels >100 pg/mL corresponded to a future ALSFRS-R slope of ∼1.5 points/month (i.e., fast progression). In the range from 40 to 100 pg/mL, ALSFRS-R slope declines quickly for each incremental increase in baseline serum NfL concentration.

## Discussion

This study comprehensively evaluated leading biochemical *prognostic* biomarker candidates, alone and in combination, and examined their potential utility when combined with established and emerging clinical predictors. This multivariate approach is essential to achieving a fuller understanding of the practical value of candidate prognostic markers. Moreover, mindful that observational studies typically enroll slower progressing patients, for greatest relevance to the design and analysis of future trials we *a priori* focused our analysis on a trial-like population, the subset of PGB participants who met clinical trial eligibility criteria. Absent a similar biomarker study that utilizes the placebo group from clinical trial(s), our approach is the most robust to date in providing clear answers about the utility of an array of prognostic biomarker candidates.

Serum NfL is a robust predictor of disease progression, whether the outcome is ASLFRS-R rate of decline or survival time. While the overlap in survival curves for the second and third quartiles of NfL (Figure 3c), for example, suggests limited prognostic value for NfL in the mid-range of values when predicting survival, the relationship between NfL and future rate of functional decline is steepest in the mid-range of values (Figure 1b). Moreover, not only does NfL provide greater prognostic value than the ENCALS predictor score, the combination of NfL and the ENCALS score yields more prognostic value than either NfL or ENCALS score alone. (Of note, we have not fully explored potential transformations of NfL data to optimize its performance as a prognostic marker beyond those displayed in Figure 1. Future research using fractional polynomials or regression splines might further improve the value of NfL as a prognostic ^34^. We also acknowledge that some information is lost by dividing a continuous prognostic into categories and that cut points for quartiles will vary from one dataset to another. The quartiles provided here are intended to be descriptive of potential non-linearity in associations, not to be prescriptive of future handling of such prognostics.) Serum pNfH, on the other hand, has some prognostic value for functional decline, but not survival; and in models already adjusting for clinical predictor(s) and NfL, it yielded no additional prognostic value. The prognostic utility of urinary p75^ECD^ and plasma miR-181ab are more nuanced, with p75^ECD^ yielding some sample size saving when combined with clinical predictor(s) and NfL (recognizing that this conclusion is based on incomplete baseline data for p75^ECD^ in this sample). Serum uric acid, albumin, creatinine, and CRP have no value as prognostic biomarkers irrespective of the outcome used. Similarly, baseline cognitive and behavioural impairment, based on the ECAS, does not add prognostic value.

While the greater prognostic value of blood NfL (than pNfH) may reflect a more critical role for the NfL isoform in maintaining neuroaxonal structure and function under pathological conditions, this may also reflect the better analytic performance of the blood NfL immunoassay. pNfH assays in blood are still hampered by a matrix effect and lack of appropriate binding reagents.^35,36^ Analytic considerations may also be relevant to the performance of urinary p75^ECD^, which has not yet achieved the same degree of analytic validation as NfL assays.^37^

The design of this study has both strengths and weaknesses. As an observational study rather than a clinical trial, a limitation is that the intervals between study visits were wide (and variable), requiring us to window study visits around defined time points for the repeated-measures analyses (see eTable 8). It is for this reason that we used observed times in a random slopes analyses to estimate sample size savings from incorporation of various potential prognostic biomarkers, despite the FDA’s preference for a repeated-measures approach for clinical trials where study visit windows are typically more rigidly controlled. Of note, many ALS clinical trials have historically used this approach ^38–40^. Moreover, the estimates themselves depend on the duration of follow-up available at the time of analysis and would likely differ over shorter or longer intervals. In addition, due to premature study closure (for administrative reasons between funding cycles) and some attrition, follow up data at 3-, 6-, and 12-month were available for only 85%, 80% and 52% of participants, respectively – resulting in less precise estimates of ALSFRS-R values beyond 6 months. Vital status after a participant’s last visit was ascertained based on clinic notes at some sites, with potentially more complete data collection on deaths; this leads to downward bias in estimates of absolute survival percentages but is unlikely to bias estimates of prognostic value. Strengths of this study include the *a priori* selection of a trial-like population, the rigorous attention to the quality of phenotypic data, and the multimodal analysis of putative prognostic biomarkers. Of note, our claims of prognostic utility do not imply any assumption of a causal relationship between a given prognostic and progression rate or survival.

We also acknowledge the limitations of the ALSFRS-R as an outcome measure in clinical trials, notably the fact that it is not uni-dimensional (meaning that items on the scale measure domains other than functional status) ^41^; that a one-point change can represent a variable amount of functional change depending on the question and the item ^42^, providing a rationale for reporting the domain specific sub-scores of the ALSFRS-R ^43^; and that the decline in ALSFRS-R is not linear across the entire course of disease ^44^. Notwithstanding these considerations, the ALSFRS-R is typically linear during the follow-up period encompassed by clinical trials ^45^, and remains the principal functional outcome measure used in ALS clinical trials ^46^.

While the longitudinal trajectory of a subset of the biomarkers was not the major focus of this investigation, we have observed subtle increases in NfL and pNfH over time (in contrast to the conventional wisdom that these are largely stable ^8,47–50^). Also noteworthy is the marked increase and relatively consistent trajectory of urinary p75^ECD^ (compared to NfL and pNfH), suggesting that urinary p75^ECD^ might have value as a response or monitoring biomarker.

It should be acknowledged that our evaluation of changes In biomarkers over time—and of the prognostic value of these biomarkers—has been conducted at a population (or group) level.

While statistically robust, conclusions from a population cannot necessarily be extrapolated to individual patients. NfL and other biomarkers considered in our analyses, therefore, remain largely research tools, with more limited (and speculative) value in the clinic setting when applied to individuals.

While confirmatory studies with larger sample sizes would add confidence to our conclusions, the results of this study are nevertheless immediately relevant to all ongoing and future ALS trials, even in the absence of formal qualification through regulatory agencies such as the FDA.^51^ Our findings are especially relevant to trials with 6-month treatment duration, the period for which we have more complete data. First, baseline NfL should be incorporated into the analysis plan for all clinical trials as a prognostic biomarker, whether functional decline or survival is used as the primary outcome. Second, how one incorporates baseline NfL into trial design – either as an eligibility criterion or as a stratification factor – depends on the purpose.

For example, if the goal is to enrich the trial population for either faster or slower progressing patients, or to stratify randomisation based on anticipated rate of disease progression, then NfL levels above or below a defined threshold might be used. Our data suggest serum NfL thresholds of <40pg/mL for slow progressors and >100pg/mL for fast progressors. Between 40-100pg/ml, given the steep relationship between NfL increase and faster future rate of ALSFRS-R decline, multiple NfL strata may be required for randomisation (as permitted by study sample size), in order to adequately control for heterogeneity of predicted disease progression rate. (Importantly, the same threshold may not hold for predicting future survival.) Third, in a hypothetical clinical trial with ALSFRS-R slope as the outcome, except for urinary p75^ECD^, other putative prognostic biomarkers yield very little in the way of sample size saving beyond those conferred by the combination of established clinical predictor(s) and NfL. While incorporation of plasma miR-181ab in such a model does not improve prediction of future rates of ALSFRS-R decline or yield additional sample size savings, it may have some value in predicting survival.

This study exemplifies the critical importance of a multivariate approach to evaluating new prognostic markers and highlights the necessity for novel markers to demonstrate value added to existing predictors. Moreover, the implication of our finding that clinical predictors (encapsulated, for example, by the ENCALS score) and blood-based measurement of NfL are strong predictors of disease progression, is that both should be incorporated into all ongoing and future Phase 2 and Phase 3 ALS trials. Moreover, the dual use of NfL as a prognostic *and* response biomarker will aid interpretation of Phase 2 trial results and facilitate go/no-go decisions about advancing experimental agents to Phase 3. Collectively, these modifications to ALS trial design and analysis should accelerate the pace of ALS therapy development.

## Supporting information

eTables

## Contributors

MB: Study concept/design, study oversight, major role in acquisition of data, analysis/interpretation of data, access to and verification of underlying data, and drafting/revising the manuscript for content.

EAM: Analysis/interpretation of data, access to and verification of underlying data, and drafting/revising the manuscript for content.

AM: Study concept/design, major role in acquisition of data, and drafting/revising the manuscript for content.

MLR: Study concept/design, major role in acquisition of data, and drafting/revising the manuscript for content.

EH: Major role in acquisition of data and drafting/revising the manuscript for content.

VL: Major role in acquisition of data.

DR: Major role in acquisition of data.

IM: Major role in acquisition of data and drafting/revising the manuscript for content.

YC: Major role in acquisition of data.

VG: Major role in acquisition of data.

JS: Major role in acquisition of data.

JH: Major role in acquisition of data.

RR: Major role in acquisition of data and editing the manuscript for content.

CAM: Major role in generation of summary cognitive/behavioural data.

LP: Study concept/design and editing the manuscript for content.

CM: Major role in acquisition of data and editing the manuscript for content.

JW: Study concept/design, study oversight, major role in acquisition of data, analysis/interpretation of data, access to and verification of underlying data, and drafting/revising the manuscript for content.

All authors have read and approved the final version of the manuscript. CReATe Consortium PGB Investigators contributed to data collection.

## Data Sharing Statement

Following publication, de-identified participant data and a data dictionary defining each field in the dataset, will be made available following request to the corresponding authors and upon execution of a data access agreement. Additional study related documents are not available.

## Declaration of Interests

MB reports grants from the NIH (U01NS107027, U54NS092091) and the ALS Association (16-TACL-242) in support of this work. He is also an unpaid member of the Board of Trustees for the ALS Association. He has served as a consultant to Alector, Alexion, Annexon, Arrowhead, Biogen, Cartesian, Denali, Eli Lilly, Horizon, Immunovant, Novartis, Roche, Sanofi, Takeda, UCB, and UniQure.

EAM reports grants from the NIH (U01NS107027, U54NS092091). He also serves as a consultant to Annexon, Biogen, Bial Biotech, Cortexyme, Chase Therapeutics, Enterin, nQ Medical, Partner Therapeutics, Stoparkinson Healthcare, and UCB. He has served on DSMBs for NeuroSense Therapeutics, Novartis, and Sanofi.

AM reports grants from the NIH (U01NS107027). He has also provided consulting services to Roche, Pfizer, and Accure Therapeutics.

MLR reports support from grants from the NIH (U01NS107027, U54NS092091) and FightMND. EH has nothing to declare.

VL has nothing to declare. DR has nothing to declare. SS has nothing to declare. IM has nothing to declare. YC has nothing to declare.

VG currently is an employee of Biohaven Pharmaceutical Inc.

JS receives research funding from the NIH< MDA, FSHD Society, Friends of FSH Research, and FSHD Canada. He also serves as a consultant or on scientific advisory boards for Avidity, Fulcrum, Dyne, Armatus, Epic Bio, Roche, Lupin, and Entrada

JH has nothing to declare.

RR reports grants from the NIH (U54NS092091) in support of this work. She is also an unpaid member of the Medical Advisory Board of the Association for Frontotemporal Dementias (AFTD) and a paid member of the Scientific Advisory Board of the Kissick Family Foundation FTD Grant Program CAM reports funding from the ALS Association and the NIH (U54NS092091).

LP reports support from the Mayo Clinic Foundation and grants from the National Institute on Aging (5P30AG0062677, U19AG063911) the National Institute of Neurological Disorders and Stroke (U54NS123743, R35NS097273, P01NS084974) and Target ALS Foundation.

CM reports grants from the NIH (AG066597, AG076411, AG066152, AG072979, NS109260, NS092091), Department of Defense, and support from the Penn Institute on Aging, Decrane Family PPA Fund, and Newhouse Fund.2

JW reports funding from the NIH (U01NS107027, U54NS092091) and the ALS Association (16-TACL-242) in support of this work.

The CReATe Consortium (U54NS092091) is part of the NIH Rare Diseases Clinical Research Network (RDCRN), an initiative of the Office of Rare Diseases Research (ORDR), NCATS. This Consortium is funded through a collaboration between NCATS and the NINDS. This work was also supported by a Clinical Trial Readiness grant (U01NS107027) from NINDS and by a grant from the ALS Association to support the CReATe Biorepository (grant ID 16-TACL-242).

## Data Availability

Following publication, de-identified participant data and a data dictionary defining each field in the dataset, will be made available following request to the corresponding authors and upon execution of a data access agreement.

## Acknowledgements

The authors thank participants in the CReATe Consortium’s Phenotype-Genotype-Biomarker (PGB1) study; research staff at each clinical site; and the CReATe Consortium project management and data management teams, genomics sub-core, and biorepository. The authors also thank members of the University of Miami Laboratory for Clinical and Biological Studies for assistance with clinical chemistry assays.

