## Supplementary material for "Prognostic Clinical and Biological Markers for Amyotrophic Lateral Sclerosis Disease Progression: Validation and Implications for Clinical Trial Design and Analysis": eTables

**Supplemental Data Tables**

**eTable 1. Baseline biomarker data, by C9orf72 repeat expansion status**

| **Biomarker** | ***C9orf72* negative** | | | ***C9orf72* positive** | | |
| --- | --- | --- | --- | --- | --- | --- |
|  | **Mean ± SD** | **Median (Q1, Q3)** | **Range** | **Mean ± SD** | **Median (Q1, Q3)** | **Range** |
| Serum NfL (pg/mL) | 73.5 ± 48.0 | 66.6 (36.2, 94.7) | 9.1 - 214.4 | 77.6 ± 37.2 | 77.9 (48.7, 90.1) | 36.3 - 200.1 |
| Serum pNfH (pg/mL) | 597 ± 718 | 267 (109, 925) | 3.4 - 4177 | 610 ± 733 | 259 (135, 844) | 21.2 - 2463 |
| Serum NfL/pNfH ratio | 0.51 ± 1.62 | 0.19 (0.086, 0.482) | 0.020 - 20.9 | 0.54 ± 0.85 | 0.16 (0.089, 0.566) | 0.041 - 3.73 |
| Urinary p75^ECD^ (ng/mg creatinine) | 5.48 ± 2.45 | 4.97 (3.84, 6.35) | 1.53 - 16.2 | 6.11 ± 2.12 | 5.54 (4.56, 7.57) | 2.71 - 10.05 |
| Serum uric acid (mg/dL) | 5.24 ± 1.32 | 5.10 (4.20, 6.20) | 2.60 - 8.90 | 4.69 ± 1.11 | 4.65 (3.95, 5.20) | 3.00 - 7.10 |
| Serum creatinine (mg/dL) | 0.78 ± 0.20 | 0.77 (0.65, 0.90) | 0.29 - 1.59 | 0.77 ± 0.14 | 0.80 (0.66, 0.88) | 0.51 - 1.04 |
| Serum albumin (g/dL) | 4.56 ± 0.34 | 4.50 (4.30, 4.80) | 3.60 - 5.80 | 4.46 ± 0.31 | 4.50 (4.30, 4.70) | 3.60 - 4.80 |
| Serum CRP (mg/dL) | 0.26 ± 0.35 | 0.10 (0.10, 0.30) | 0.10 - 2.30 | 0.28 ± 0.29 | 0.10 (0.10, 0.40) | 0.10 - 0.90 |
| Plasma miR-181a (UMI) | 454 ± 209 | 419 (316, 554) | 124 - 1699 | 425 ± 199 | 392 (300, 474) | 170 - 962 |
| Plasma miR-181b (UMI) | 67.5 ± 32.3 | 63.6 (44.9, 80.5) | 13.8 - 263 | 54.3 ± 19.5 | 54.1 (41.3, 58.8) | 24.7 - 107.5 |
| Plasma miR-181ab (UMI^2) | 35731 ± 40867 | 26288 (14240, 43215) | 3148 - 447480 | 25001 ± 19655 | 20136 (12350, 26769) | 6661 - 88047 |

**eTable 2. Baseline biomarker data, by sex**

| **Biomarker** | **Female** | | | **Male** | | |
| --- | --- | --- | --- | --- | --- | --- |
|  | **Mean ± SD** | **Median (Q1, Q3)** | **Range** | **Mean ± SD** | **Median (Q1, Q3)** | **Range** |
| Serum NfL (pg/mL) | 89.0 ± 52.8 | 76.9 (44.8, 121.5) | 11.0 - 214.4 | 61.7 ± 37.8 | 57.6 (33.3, 78.8) | 9.1 - 186.3 |
| Serum pNfH (pg/mL) | 650 ± 730 | 355 (118, 968) | 3.4 - 4177 | 556 ± 709 | 218 (102, 725) | 8.0 - 3555 |
| Serum NfL/pNfH ratio | 0.67 ± 2.25 | 0.17 (0.093, 0.482) | 0.027 - 20.89 | 0.39 ± 0.55 | 0.20 (0.078, 0.505) | 0.020 - 3.73 |
| Urinary p75^ECD^ (ng/mg creatinine) | 5.34 ± 2.57 | 4.71 (3.59, 6.17) | 1.53 - 16.15 | 5.68 ± 2.31 | 5.30 (4.14, 6.57) | 1.62 - 13.8 |
| Serum uric acid (mg/dL) | 4.49 ± 1.12 | 4.40 (3.60, 5.00) | 2.60 - 7.90 | 5.76 ± 1.17 | 5.80 (4.90, 6.60) | 3.20 - 8.90 |
| Serum creatinine (mg/dL) | 0.70 ± 0.17 | 0.68 (0.59, 0.82) | 0.37 - 1.36 | 0.85 ± 0.20 | 0.84 (0.74, 0.94) | 0.29 - 1.59 |
| Serum albumin (g/dL) | 4.45 ± 0.34 | 4.50 (4.20, 4.70) | 3.60 - 5.50 | 4.63 ± 0.32 | 4.60 (4.40, 4.80) | 3.80 - 5.80 |
| Serum C-reactive protein (mg/dL) | 0.32 ± 0.40 | 0.10 (0.10, 0.40) | 0.10 - 2.30 | 0.22 ± 0.29 | 0.10 (0.10, 0.10) | 0.10 - 2.00 |
| Plasma miR-181a (UMI) | 432 ± 177 | 415 (310, 540) | 124 - 916 | 467 ± 229 | 421 (316, 564) | 198 - 1699 |
| Plasma miR-181b (UMI) | 65.2 ± 28.7 | 60.2 (43.1, 78.8) | 13.8 - 150.1 | 67.0 ± 33.7 | 63.1 (46.0, 80.2) | 13.8 - 263.4 |
| Plasma miR-181ab (UMI^2) | 31161 ± 24248 | 23956 (13470, 42694) | 3225 - 108896 | 37502 ± 48214 | 26288 (13638, 42977) | 3148 - 447480 |

**eTable 3. Longitudinal biomarker trajectories, by sex**

| **Biomarker** | **Increase per month, relative to baseline** | | | | |
| --- | --- | --- | --- | --- | --- |
|  | **Female** | | **Male** | | **p-value** |
|  | **Mean** | **(95% CI)** | **Mean** | **(95% CI)** |  |
| Serum NfL (pg/mL) | 1.40% | (0.79%, 2.02%) | 0.57% | (0.06%, 1.09%) | 0.043 |
| Serum pNfH (pg/mL) | 0.40% | (-0.49%, 1.30%) | 0.50% | (-0.26%, 1.26%) | 0.87 |
| Serum NfL/pNfH ratio | 1.02% | (0.21%, 1.84%) | 0.03% | (-0.64%, 0.71%) | 0.068 |
| Urinary p75^ECD^ (ng/mg creatinine) | 2.86% | (1.89%, 3.84%) | 2.45% | (1.73%, 3.18%) | 0.51 |

**eTable 4. Correlation matrix showing relationship between individual components of ENCALS prediction score and serum NfL**

|  | **Age**  **at onset** | **Bulbar symptoms**  **at onset** | **Diagnostic**  **delay** | **Baseline**  **El Escorial**  **definite** | **Baseline**  **ΔFRS** | **Baseline SVC**  **%predicted** | **C9orf72**  **positive** | **Baseline**  **serum NfL** |
| --- | --- | --- | --- | --- | --- | --- | --- | --- |
| **Age**  **at onset** | -- |  |  |  |  |  |  |  |
| **Bulbar symptoms**  **at onset** | 0.123  (-0.015, 0.256) p=0.081 | -- |  |  |  |  |  |  |
| **Diagnostic**  **delay** | 0.035  (-0.103, 0.172) 0.62 | -0.041  (-0.177, 0.098) 0.56 | -- |  |  |  |  |  |
| **Baseline**  **El Escorial**  **definite** | 0.083  (-0.055, 0.218) 0.24 | 0.201  (0.065, 0.329) 0.0039 | 0.047  (-0.091, 0.184) 0.51 | -- |  |  |  |  |
| **Baseline**  **ΔFRS** | 0.187  (0.051, 0.317) 0.0072 | 0.142  (0.004, 0.274) 0.043 | -0.426  (-0.532, -0.306) <0.0001 | 0.166  (0.029, 0.297) 0.017 | -- |  |  |  |
| **Baseline SVC**  **%predicted** | -0.033  (-0.170, 0.105) 0.64 | -0.112  (-0.245, 0.027) 0.11 | 0.006  (-0.132, 0.143) 0.93 | -0.144  (-0.277, -0.007) 0.039 | -0.164  (-0.295, -0.027) 0.019 | -- |  |  |
| **C9orf72**  **positive** | -0.063  (-0.199, 0.075) 0.37 | 0.055  (-0.084, 0.191) 0.44 | 0.055  (-0.084, 0.191) 0.44 | 0.022  (-0.116, 0.160) 0.75 | 0.117  (-0.021, 0.251) 0.10 | -0.032  (-0.169, 0.106) 0.65 | -- |  |
| **Baseline**  **serum NfL** | 0.275  (0.143, 0.398) <0.0001 | 0.234  (0.100, 0.360) 0.0007 | -0.078  (-0.214, 0.060) 0.27 | 0.222  (0.087, 0.349) 0.0014 | 0.279  (0.147, 0.401) <0.0001 | -0.027  (-0.164, 0.110) 0.70 | 0.026  (-0.112, 0.163) 0.71 | -- |

Pearson correlation coefficients (95% confidence intervals), and associated p-values

**eTable 5. Prognostic markers of survival: Linear effect per SD**

Estimates are the modelled hazard ratio (95% confidence interval) for a 1 standard deviation increase in the prognostic marker.

| **Prognostic marker** | **Unadjusted** | **Covariate(s) included** | | | |
| --- | --- | --- | --- | --- | --- |
|  |  | **Core clinical predictors ^1^** | **ENCALS predictor score** | **Core clinical predictors ^1^**  **& NfL** | **ENCALS predictor score**  **& NfL** |
| Sex, male | 0.95 (0.77, 1.17) | 1.06 (0.84, 1.34) | 0.99 (0.80, 1.22) | 1.20 (0.95, 1.54) | 1.17 (0.94, 1.46) |
| Age at onset, years | 1.40 (1.13, 1.75) | 0.51 (0.01, 44.5) | 1.05 (0.81, 1.35) | 0.66 (0.01, 68.6) | 0.93 (0.71, 1.22) |
| Bulbar symptoms at onset | 1.23 (1.00, 1.49) | -- | 1.13 (0.92, 1.38) | -- | 0.98 (0.79, 1.20) |
| Diagnostic delay | 0.78 (0.61, 0.97) | -- | 1.43 (1.02, 2.00) | -- | 1.08 (0.77, 1.52) |
| Baseline ΔFRS | 1.61 (1.36, 1.89) | -- | 1.18 (0.91, 1.51) | -- | 1.15 (0.87, 1.47) |
| Baseline age | 1.38 (1.12, 1.73) | -- | 1.05 (0.82, 1.35) | -- | 0.93 (0.71, 1.21) |
| Baseline ALSFRS-R | 0.70 (0.58, 0.85) | 0.79 (0.63, 1.02) | 0.84 (0.68, 1.04) | 0.82 (0.64, 1.07) | 0.88 (0.71, 1.10) |
| Baseline SVC %predicted | 0.72 (0.58, 0.89) | 0.80 (0.64, 0.99) | 0.89 (0.71, 1.10) | 0.74 (0.60, 0.91) | 0.82 (0.66, 1.01) |
| Baseline ECAS total | 1.02 (0.84, 1.28) | 1.14 (0.90, 1.48) | 1.03 (0.82, 1.33) | 1.12 (0.89, 1.43) | 1.06 (0.86, 1.35) |
| Baseline ECAS ALS-specific | 1.05 (0.85, 1.34) | 1.19 (0.93, 1.56) | 1.07 (0.84, 1.39) | 1.17 (0.92, 1.53) | 1.12 (0.89, 1.45) |
| Baseline ECAS ALS non-specific | 0.95 (0.79, 1.19) | 1.00 (0.81, 1.26) | 0.92 (0.75, 1.17) | 0.98 (0.80, 1.22) | 0.94 (0.78, 1.16) |
| Baseline cognitive impairment | 1.00 (0.75, 1.27) | 0.89 (0.66, 1.15) | 0.94 (0.71, 1.20) | 1.02 (0.74, 1.34) | 1.07 (0.80, 1.38) |
| Baseline behavioural impairment | 1.11 (0.80, 1.45) | 1.02 (0.72, 1.37) | 1.20 (0.87, 1.58) | 0.97 (0.68, 1.31) | 1.12 (0.81, 1.49) |
| Baseline ENCALS predictor score | 2.35 (1.74, 3.21) | 2.74 (1.36, 5.96) | -- | 2.23 (1.18, 4.59) | -- |
| Baseline serum NfL | 1.97 (1.58, 2.48) | 1.86 (1.45, 2.41) | 1.83 (1.45, 2.33) | -- | -- |
| Baseline serum pNfH | 1.27 (1.05, 1.55) | 1.23 (1.00, 1.52) | 1.24 (1.01, 1.53) | 0.94 (0.76, 1.18) | 0.91 (0.73, 1.15) |
| Baseline urinary p75^ECD^ | 0.99 (0.79, 1.24) | 0.93 (0.71, 1.19) | 0.85 (0.66, 1.09) | 0.92 (0.71, 1.18) | 0.89 (0.69, 1.14) |
| Baseline serum uric acid | 0.88 (0.72, 1.06) | 0.96 (0.78, 1.18) | 0.92 (0.76, 1.12) | 1.04 (0.85, 1.28) | 1.01 (0.83, 1.23) |
| Baseline serum creatinine | 1.05 (0.87, 1.29) | 1.05 (0.85, 1.30) | 1.06 (0.87, 1.30) | 1.01 (0.82, 1.26) | 1.00 (0.81, 1.24) |
| Baseline serum albumin | 0.88 (0.72, 1.07) | 0.95 (0.78, 1.17) | 0.96 (0.79, 1.18) | 0.96 (0.78, 1.18) | 0.98 (0.80, 1.21) |
| Baseline serum CRP | 0.94 (0.74, 1.16) | 1.02 (0.79, 1.29) | 1.03 (0.81, 1.28) | 0.96 (0.75, 1.22) | 0.92 (0.72, 1.15) |
| Baseline plasma miR-181ab | 1.36 (1.09, 1.69) | 1.36 (1.08, 1.72) | 1.33 (1.06, 1.67) | 1.36 (1.08, 1.71) | 1.32 (1.06, 1.64) |
| Baseline miR-181ab > 39,300 UMI ^2^ | 1.32 (1.07, 1.61) | 1.22 (0.98, 1.51) | 1.22 (0.99, 1.49) | 1.24 (1.00, 1.54) | 1.22 (1.00, 1.50) |
| Baseline miR-181ab > 24,590 UMI ^3^ | 1.30 (1.05, 1.61) | 1.28 (1.03, 1.61) | 1.31 (1.06, 1.64) | 1.37 (1.09, 1.75) | 1.36 (1.09, 1.70) |
| Baseline NfL+miR181ab poor Px ^4^ | 1.67 (1.36, 2.03) | 1.58 (1.25, 2.00) | 1.56 (1.27, 1.92) | 1.14 (0.84, 1.53) | 1.14 (0.86, 1.50) |
| Baseline NfL+miR181ab poor Px ^5^ | 1.90 (1.53, 2.38) | 1.77 (1.40, 2.26) | 1.79 (1.43, 2.25) | 1.34 (0.97, 1.87) | 1.39 (1.01, 1.93) |
| Baseline NfL median split ^6^ | 1.67 (1.35, 2.08) | 1.50 (1.19, 1.90) | 1.52 (1.22, 1.89) | 0.90 (0.63, 1.28) | 0.92 (0.65, 1.31) |
| Baseline NfL 4-level split ^7^ | 1.89 (1.53, 2.36) | 1.73 (1.37, 2.20) | 1.71 (1.37, 2.15) | 1.09 (0.66, 1.80) | 1.08 (0.65, 1.77) |

^1^ Core clinical predictors in survival analyses include bulbar onset, diagnostic delay, ΔFRS, and baseline age.

^2^ Threshold of 39,300 UMI in plasma as defined by Magen et al ^11^.

^3^ Median of 24,590 UMI in plasma in the current dataset.

^4^ Poor prognosis based on published optimal combination of NfL and miR-181ab, in which a poor prognostic factor is defined as either (NfL > 109.8 pg/mL) or (NfL > 59.0 pg/mL and miR-181ab > 39,300 UMI) ^11^.

^5^ Poor prognosis based on recalculated combination of NfL and mIR-181ab using thresholds obtained from the current dataset; a poor prognostic factor is defined as either (NfL > 80.8 pg/mL) or (NfL > 44.8 pg/mL and mIR-181ab > 24,590 UMI).

^6^ Median serum NfL = 67.9 pg/mL.

^7^ Serum NfL 4-level split is at the 33^rd^, 50^th^, and 67^th^ percentiles (44.8 pg/mL, 67.9 pg/mL, and 80.8 pg/mL, respectively), i.e., tertiles and median, rather than quartiles, to mimic construction of the NfL+miR18ab measure ^11^.

**eTable 6. Prognostic markers of functional decline: Linear effect per SD**

Estimates are the linear predictors of ALSFRS-R slope (95% confidence interval) for a 1 standard deviation increase in the prognostic marker.

| **Prognostic Marker** | **Unadjusted** | **Covariate(s) included** | | | |
| --- | --- | --- | --- | --- | --- |
|  |  | **Core clinical predictors ^1^** | **ENCALS predictor score** | **Core clinical predictors ^1^**  **& NfL** | **ENCALS predictor score**  **& NfL** |
| Sex, male | 0.014 (-0.09, 0.121) | -0.020 (-0.13, 0.091) | 0.011 (-0.09, 0.114) | -0.13 (-0.22, -0.03) | -0.12 (-0.21, -0.03) |
| Age at onset, years | -0.099 (-0.21, 0.007) | -0.063 (-0.17, 0.040) | 0.039 (-0.08, 0.160) | -0.002 (-0.09, 0.088) | 0.087 (-0.02, 0.191) |
| Bulbar symptoms at onset | -0.11 (-0.22, -0.01) | -- | -0.079 (-0.18, 0.025) | -- | 0.0021 (-0.09, 0.096) |
| Diagnostic delay | 0.16 (0.060, 0.269) | -- | 0.059 (-0.06, 0.179) | -- | 0.096 (-0.01, 0.199) |
| Baseline ΔFRS | -0.22 (-0.33, -0.11) | -- | -0.13 (-0.26, 0.003) | -- | -0.072 (-0.19, 0.045) |
| Age at baseline | -0.089 (-0.20, 0.017) | -0.062 (-0.17, 0.041) | 0.044 (-0.07, 0.163) | -0.002 (-0.09, 0.087) | 0.089 (-0.01, 0.190) |
| Baseline ALSFRS-R | 0.040 (-0.08, 0.156) | -0.081 (-0.22, 0.061) | -0.030 (-0.15, 0.086) | -0.092 (-0.22, 0.031) | -0.061 (-0.16, 0.039) |
| Baseline SVC %predicted | 0.13 (0.023, 0.242) | 0.095 (-0.01, 0.202) | 0.083 (-0.02, 0.192) | 0.11 (0.014, 0.197) | 0.094 (0.001, 0.188) |
| Baseline ECAS total | 0.037 (-0.08, 0.153) | 0.024 (-0.09, 0.137) | 0.021 (-0.09, 0.132) | 0.020 (-0.08, 0.119) | 0.0008 (-0.10, 0.098) |
| Baseline ECAS ALS-specific | 0.033 (-0.08, 0.149) | 0.020 (-0.09, 0.133) | 0.021 (-0.09, 0.132) | 0.012 (-0.09, 0.112) | -0.002 (-0.10, 0.096) |
| Baseline ECAS ALS non-specific | 0.033 (-0.08, 0.151) | 0.026 (-0.09, 0.141) | 0.012 (-0.10, 0.125) | 0.032 (-0.07, 0.133) | 0.0081 (-0.09, 0.107) |
| Baseline cognitive impairment | -0.021 (-0.15, 0.102) | -0.016 (-0.14, 0.105) | -0.019 (-0.14, 0.101) | -0.037 (-0.15, 0.072) | -0.031 (-0.14, 0.076) |
| Baseline behavioural impairment | -0.094 (-0.24, 0.052) | -0.067 (-0.21, 0.077) | -0.075 (-0.21, 0.064) | 0.011 (-0.11, 0.136) | 0.018 (-0.10, 0.137) |
| Baseline ENCALS predictor score | -0.22 (-0.32, -0.12) | -0.12 (-0.25, 0.013) | -- | -0.052 (-0.17, 0.062) | -- |
| Baseline serum NfL | -0.41 (-0.50, -0.31) | -0.38 (-0.48, -0.28) | -0.37 (-0.46, -0.28) | -- | -- |
| Baseline serum pNfH | -0.16 (-0.27, -0.06) | -0.15 (-0.26, -0.05) | -0.15 (-0.25--0.04) | 0.053 (-0.05, 0.159) | 0.061 (-0.04, 0.167) |
| Baseline urinary p75^ECD^ | -0.019 (-0.14, 0.097) | -0.020 (-0.13, 0.094) | 0.0081 (-0.10, 0.119) | 0.0022 (-0.09, 0.099) | 0.014 (-0.08, 0.110) |
| Baseline serum uric acid | 0.062 (-0.05, 0.170) | 0.032 (-0.07, 0.136) | 0.053 (-0.05, 0.156) | -0.028 (-0.12, 0.062) | -0.020 (-0.11, 0.071) |
| Baseline serum creatinine | -0.008 (-0.11, 0.098) | -0.012 (-0.11, 0.088) | 0.0047 (-0.10, 0.105) | -0.009 (-0.10, 0.077) | 0.0012 (-0.09, 0.088) |
| Baseline serum albumin | 0.0086 (-0.10, 0.119) | -0.038 (-0.14, 0.069) | -0.042 (-0.15, 0.066) | -0.058 (-0.15, 0.034) | -0.066 (-0.16, 0.026) |
| Baseline serum CRP | -0.066 (-0.18, 0.044) | -0.092 (-0.20, 0.015) | -0.075 (-0.18, 0.031) | -0.055 (-0.15, 0.039) | -0.033 (-0.13, 0.060) |
| Baseline plasma miR-181ab | -0.14 (-0.25, -0.03) | -0.11 (-0.21, -0.00) | -0.13 (-0.23, -0.03) | -0.084 (-0.18, 0.009) | -0.099 (-0.19, -0.01) |
| Baseline miR-181ab > 39,300 UMI ^2^ | -0.13 (-0.24, -0.02) | -0.10 (-0.21, 0.006) | -0.11 (-0.21, -0.01) | -0.082 (-0.18, 0.011) | -0.087 (-0.18, 0.002) |
| Baseline miR-181ab > 24,590 UMI ^3^ | -0.12 (-0.23, -0.02) | -0.091 (-0.20, 0.015) | -0.12 (-0.22, -0.02) | -0.068 (-0.16, 0.023) | -0.085 (-0.17, 0.003) |
| Baseline NfL+miR181ab poor Px ^4^ | -0.33 (-0.43, -0.23) | -0.30 (-0.41, -0.19) | -0.29 (-0.39, -0.19) | -0.095 (-0.22, 0.032) | -0.083 (-0.21, 0.042) |
| Baseline NfL+miR181ab poor Px ^5^ | -0.36 (-0.46, -0.26) | -0.32 (-0.42, -0.23) | -0.32 (-0.42, -0.23) | -0.13 (-0.26, 0.000) | -0.13 (-0.26, 0.000) |
| Baseline NfL median split ^6^ | -0.37 (-0.47, -0.28) | -0.34 (-0.43, -0.24) | -0.33 (-0.43, -0.24) | -0.13 (-0.28, 0.010) | -0.13 (-0.28, 0.011) |
| Baseline NfL 4-level split ^7^ | -0.40 (-0.50, -0.31) | -0.38 (-0.47, -0.28) | -0.37 (-0.46, -0.27) | -0.21 (-0.41, -0.01) | -0.19 (-0.39, 0.009) |

^1^ Core clinical predictors in functional decline analyses include bulbar onset, diagnostic delay, and ΔFRS.

^2^ Threshold of 39,300 UMI in plasma as defined by Magen et al ^11^.

^3^ Median of 24,590 UMI in plasma in the current dataset.

^4^ Poor prognosis based on published optimal combination of NfL and miR-181ab, in which a poor prognostic factor is defined as either (NfL > 109.8 pg/mL) or (NfL > 59.0 pg/mL and miR-181ab > 39,300 UMI) ^11^.

^5^ Poor prognosis based on recalculated combination of NfL and mIR-181ab using thresholds obtained from the current dataset; a poor prognostic factor is defined as either (NfL > 80.8 pg/mL) or (NfL > 44.8 pg/mL and mIR-181ab > 24,590 UMI).

^6^ Median serum NfL = 67.9 pg/mL.

^7^ Serum NfL 4-level split is at the 33^rd^, 50^th^, and 67^th^ percentiles (44.8 pg/mL, 67.9 pg/mL, and 80.8 pg/mL, respectively), i.e., tertiles and median, rather than quartiles, to mimic construction of the NfL+miR18ab measure ^11^.

**eTable 7. Multiplicity-adjusted p-values for prognostic markers of survival (Table 3) and functional decline (Table 4)**

| **Prognostic Marker** | **Table 3** | | | **Table 4** | |
| --- | --- | --- | --- | --- | --- |
|  | **Unadjusted** | **Core clinical**  **adjusted model ^1^** | **ENCALS**  **adjusted model ^2^** | **Unadjusted** | **ENCALS**  **adjusted model ^2^** |
| Sex, male | 1 | 1 | 1 | 1 | 1 |
| Age at onset, years | 0.37 | 1 | 1 | 1 | 1 |
| Bulbar symptoms at onset | 0.59 | (n/a) | 1 | 0.61 | 1 |
| Diagnostic delay | 1 | 1 | 1 | 0.35 | 1 |
| Baseline ΔFRS | <0.0001 | 1 | 1 | 0.0005 | 0.083 |
| Baseline age | 1 | 1 | 1 | 1 | 1 |
| Baseline ALSFRS-R | 0.020 | 0.67 | 0.96 | 1 | 1 |
| Baseline SVC %predicted | 0.093 | 1 | 1 | 1 | 1 |
| Baseline ECAS total | 1 | 1 | 1 | 1 | 1 |
| Baseline ECAS ALS-specific | 1 | 1 | 1 | 1 | 1 |
| Baseline ECAS ALS-non-specific | 1 | 1 | 1 | 1 | 1 |
| Baseline cognitive impairment | 1 | 1 | 1 | 1 | 1 |
| Baseline behavioural impairment | 1 | 1 | 1 | 1 | 1 |
| Baseline ENCALS predictor score | <0.0001 | 1 | 1 | 0.0001 | 0.46 |
| Baseline serum NfL | <0.0001 | <0.0001 | <0.0001 | <0.0001 | <0.0001 |
| Baseline serum pNfH | 1 | 1 | 1 | 0.43 | 0.75 |
| Baseline urinary p75^ECD^ | 1 | 1 | 1 | 1 | 1 |
| Baseline serum uric acid | 1 | 1 | 1 | 1 | 1 |
| Baseline serum creatinine | 1 | 1 | 1 | 1 | 1 |
| Baseline serum albumin | 1 | 1 | 1 | 1 | 0.75 |
| Baseline serum CRP | >0.99 | >0.99 | >0.99 | >0.99 | >0.99 |
| Baseline plasma miR-181ab | 0.35 | >0.99 | >0.99 | 0.98 | >0.99 |
| Baseline miR-181ab > 39,300 UMI ^3^ | 0.12 | >0.99 | >0.99 | 0.35 | 0.75 |
| Baseline miR-181ab > 24,590 UMI ^4^ | 0.28 | 0.67 | 0.32 | 0.43 | 0.46 |
| Baseline NfL+miR181ab poor Px ^5^ | <0.0001 | 0.0033 | 0.0007 | <0.0001 | <0.0001 |
| Baseline NfL+miR181ab poor Px ^6^ | <0.0001 | <0.0001 | <0.0001 | <0.0001 | <0.0001 |
| Baseline NfL median split ^7^ | <0.0001 | 0.015 | 0.0046 | <0.0001 | <0.0001 |
| Baseline NfL 4-level split ^8^ | <0.0001 | 0.0013 | 0.0009 | <0.0001 | <0.0001 |

^1^ Core clinical predictors in survival analyses include bulbar onset, diagnostic delay, ΔFRS, and baseline age.

^2^ ENCALS predictor score is derived from ΔFRS, bulbar onset, diagnostic delay, age at onset, SVC percent predicted, El Escorial definite ALS, presence of FTD, and presence of a *C9orf72* repeat expansion.

^3^ Threshold of 39,300 UMI in plasma as defined by Magen et al ^11^.

^4^ Median of 24,590 UMI in plasma in the current dataset.

^5^ Poor prognosis based on published optimal combination of NfL and miR-181ab, in which a poor prognostic factor is defined as either (NfL > 109.8pg/ml) or (NfL > 59.0pg/ml and miR-181ab > 39,300 UMI) ^11^.

^6^ Poor prognosis based on recalculated combination of NfL and miR-181ab using thresholds obtained from the current dataset; a poor prognostic factor is defined as either (NfL > 80.8 pg/mL) or (NfL > 44.8 pg/mL and mIR-181ab > 24,590 UMI).

^7^ Median serum NfL = 67.9 pg/mL.

^8^ Serum NfL 4-level split is at the 33^rd^, 50^th^, and 67^th^ percentiles (44.8 pg/mL, 67.9 pg/mL, and 80.8 pg/mL, respectively), i.e., tertiles and median, rather than quartiles, to mimic construction of the NfL+miR18ab measure ^11^.

**eTable 8. Windowed visits for mixed model repeated-measures analyses**

| **Windowed**  **Visit** | **N** | **Mean (SD) of**  **Follow-up Duration** |
| --- | --- | --- |
| Month 0 | 203 | 0 |
| Month 3 | 173 | 3.13 (0.63) |
| Month 6 | 162 | 6.54 (1.09) |
| Month 12 | 106 | 12.07 (1.43) |
| Month 18 | 51 | 17.95 (1.70) |
